# Roles of community and sexual contacts as drivers of clade I mpox outbreaks

**DOI:** 10.1101/2024.10.15.24315554

**Authors:** Hiroaki Murayama, Toshiaki R. Asakura, Borame L. Dickens, Dorothy Boyle, Jen Han Foo, Shihui Jin, Patrick K. Mukadi, Keisuke Ejima, Sung-mok Jung, Akihiro Nishi, Kiesha Prem, Audry M. Wakamba, Diafuka Saila-Ngita, David Niyukuri, Akira Endo

**Affiliations:** School of Medicine, International University of Health and Welfare, Narita, Japan; Department of Infectious Disease Epidemiology, London School of Hygiene & Tropical Medicine, London, UK; Centre of Mathematical Modelling of Infectious Diseases, London School of Hygiene & Tropical Medicine, London, UK; School of Tropical Medicine and Global Health, Nagasaki University, Nagasaki, Japan; Saw Swee Hock School of Public Health, National University of Singapore, Singapore; Graduate School of Biomedical Sciences, Nagasaki University, Nagasaki, Japan; Department of Clinical Infectious Diseases, Institute of Tropical Medicine, Nagasaki University, Nagasaki, Japan; Department of Epidemiology and Global Health, Institut de Recherche Biomédicale, Kinshasa, Democratic Republic of Congo; Lee Kong Chian School of Medicine, Nanyang Technological University, Singapore, Singapore; National Centre for Infectious Diseases, Singapore, Singapore; Carolina Population Center, University of North Carolina at Chapel Hill, Chapel Hill, NC, USA; Department of Epidemiology, Fielding School of Public Health, University of California, Los Angeles, CA, USA; California Center for Population Research, University of California, Los Angeles, CA, USA; Ministry of Health, National Expanded Program for Immunization, Kinshasa, DRC; Department of Basic Sciences, Faculty of Veterinary Medicine, Université de Kinshasa (UNIKIN), Kinshasa, DRC; Doctoral School, University of Burundi, Bujumbura, Burundi; Department of Mathematics, University of Burundi, Bujumbura, Burundi; The South African Department of Science and Technology-National Research Foundation (DST-NRF) Centre of Excellence in Epidemiological Modelling and Analysis (SACEMA), Stellenbosch University, Cape Town, South Africa

## Abstract

Initial investigation into the emerging mpox outbreak of novel clade Ib in eastern Democratic Republic of the Congo has identified signs of sustained human-to-human transmission and epidemiological links to sexual contacts involving female sex workers (FSWs)^1^, which have not been observed in previous clade Ia outbreaks. Using mathematical models incorporating age-dependent contact patterns, we quantified the role of frequent sexual interactions as opposed to community contacts in clade Ib’s dynamics and found that this additional mode of transmission could explain its increased outbreak potential compared with clade Ia. As with the globally-circulating clade IIb, transmitted predominantly among men who have sex with men^2^, our findings reinforce the importance of protecting key population groups, specifically FSWs for clade Ib, in controlling ongoing mpox outbreaks.

## Main text

Since September 2023, a sustained local transmission of a novel subclade of monkeypox virus (MPXV), classified as clade Ib, has been observed in the Democratic Republic of the Congo (DRC) along with its spread to multiple other countries^3^. In response to the escalating situation, the World Health Organization (WHO) declared a public health emergency of international concern (PHEIC) on 14 August 2024^4^. Clade Ib was first identified from a cluster of cases in the Kamituga health zone of the South Kivu province, eastern DRC, where an initial investigation found epidemiological links to sexual contacts and a significant proportion of female sex workers (FSWs) among reported cases^5^. Genetic analyses have indicated a distinct profile from clade Ia circulating in other parts of DRC, characterised by the accumulation of apolipoprotein B mRNA editing enzyme catalytic subunit 3 (APOBEC3)-associated mutations^5^. Those mutations suggest sustained human-to-human transmission and were also observed in the globally-circulating MPXV clade IIb, which primarily affected men who have sex with men (MSM)^2^. Over the summer of 2024, clade Ib showed steady growth within many health zones in the North and South Kivu provinces and introductions to other provinces of DRC^6^. Additionally, international spread to neighbouring countries (Burundi, Uganda, Rwanda, Kenya, Zambia and Zimbabwe) and beyond the African continent (Sweden, Thailand, India, Germany and UK) had been confirmed by October 2024^7^, of which Burundi had reported the largest number of cases (1,509 as of 27 October 2024^8^). Thereafter, the mpox outbreak has continued to expand predominantly within Africa, with nearly 20,000 cases in 2025 alone as of May^9^. The number of countries ever reporting clade Ib cases have increased to 12 within Africa and 29 globally, including sporadic suspected local transmission events outside of Africa^9^.

MPXV clade Ia has been circulating in DRC and other Central African countries for over 50 years^10^ via zoonotic transmission followed by self-limiting chains of human-to-human transmission through close contact, mostly within households^11^. Since the cessation of the smallpox vaccination programme in 1980, which had provided cross-protection against mpox^12–17^, DRC has seen a consistent rise in suspected mpox cases^18,19^, from 3.0 per 100,000 people in 2010 to 11.5 per 100,000 people in 2023^20^. Despite this increase, zoonotic spillover remained as the primary driver of outbreaks^21,22^, and there has been no evidence of large clade I outbreaks solely sustained by human-to-human transmission before the emergence of clade Ib in 2023. In contrast, the clade Ib outbreak in the South Kivu province has shown a clear exponential growth trend, suggesting human-to-human contact as the primary mode of transmission^23,24^. The rapid rise of clade Ib cases with a potential increase in the contribution of human-to-human transmission relative to clade Ia is of growing global concern, especially given that the outbreak is not restricted to specific at-risk populations but is affecting broader groups, including children^25^.

The extensive clade Ib case profiles including both children and adults along with documented occupational risks among FSWs may reflect its mixed transmission routes through regular contacts in the community and through sexual contacts. Sexual contact as a novel route of transmission has also been documented in the ongoing global mpox outbreak caused by clade IIb since 2022 and was suggested to have contributed to the rapid spread among MSM^26,27^; meanwhile, sustained transmission of clade IIb through community or heterosexual contact routes have not been observed and cases were mostly restricted to the MSM group^28^. If clade Ib, transmissible through heterosexual contact, continues to grow into another international mpox outbreak following clade IIb, the potentially more emphasised role of community contacts in its transmission than clade IIb may enable it to affect a larger susceptible population and pose challenges for control. Although the involvement of sexual contacts among FSWs and their clients has been well documented in the initial spread of clade Ib in the Kamituga health zone in South Kivu^25^, little is currently understood on the relative contribution of community and sexual contacts to subsequent clade Ib dynamics in broader settings not limited to Kamituga, and how they may explain the distinct epidemiological features of clade Ib compared to clade Ia. With the gradual expansion of clade Ib within DRC and beyond borders, analytical efforts to identify its plausible transmission mechanisms and priority groups for prevention should leverage the best available evidence, if limited, to inform and support effective control policies.

In this study, we hypothesised that the rapid growth of the clade Ib outbreak could be explained by its entry into a key population group with a high rate of sexual contact. We developed a mathematical model for MPXV transmission dynamics incorporating both community contact route and sexual contact route involving FSWs. We first used a model incorporating only a community contact route to replicate the age profiles of the current and historical clade Ia outbreak datasets. We then expanded the model to include a sexual contact route to quantify the contribution of each route to the overall clade Ib transmission dynamics.

### Characterising community contact transmission patterns in historical and current clade Ia outbreaks

The age-dependent contact matrix, whose entries represent the average number of contacts (typically defined as either conversational^a^ or physical contacts) that an individual of a specific age group has with other age groups per unit time, is a common tool for modelling transmission dynamics of directly-transmitted infections across multiple age groups.

Assuming that the risk of transmission is proportional to the frequency of relevant types of contacts one experiences with infectious individuals, the contact matrix is associated with the so-called next generation matrix, a matrix that is multiplied by a vector containing the number of cases by age groups in one generation to produce a vector for the next generation^29^. During the exponential phase of an outbreak, the age distribution of cases would quickly converge to the dominant eigenvector of the next generation matrix and the dominant eigenvalue would define the effective reproduction number^29^. We modelled the next generation matrix by combining a contact matrix informed by empirical contact survey data from Zimbabwe^30^ and age-dependent susceptibility that we assumed to reflect potentially higher susceptibility among children aged 0–4 years^31^ and lower susceptibility among smallpox-immunised birth cohorts born in or before 1980^12–17^.

This simple model, validated by hold-out data (empirical Kullback-Leibler divergence < 0.01; Extended Data Table 2 and Figure S1), well described the age distribution for both the historical clade Ia cases in the Tshuapa province (from a well-documented clade Ia outbreak between 2011-2015^32^), DRC and the recent cases in six provinces of DRC where clade Ia has been endemic, with a decade’s shift in the age of smallpox-immunised cohorts from 34 and above to 45 and above (Figures 1A and B). Our model employed four contact matrices with different contact definitions; since all four models were overall similar in goodness-of-fit, we used model averaging^33^ to pool the model outputs (Table 1). The estimated susceptibility among children aged 0-4 (model averaging estimate: 1.5 [95% credible interval: 1.05–2.1]) and smallpox-immunised cohort (0.26 [0.17–0.38]) relative to other age groups was consistent with literature: ratios of 1.3–1.6 between the historical secondary attack risk estimates for clade Ia among age 0–4 and 5–14^31^ and the estimated effectiveness of past smallpox vaccines against clade IIb of around 70–75%^17,34^ combined with high historical coverage in DRC (89–97%)^35^

**Figure 1.**
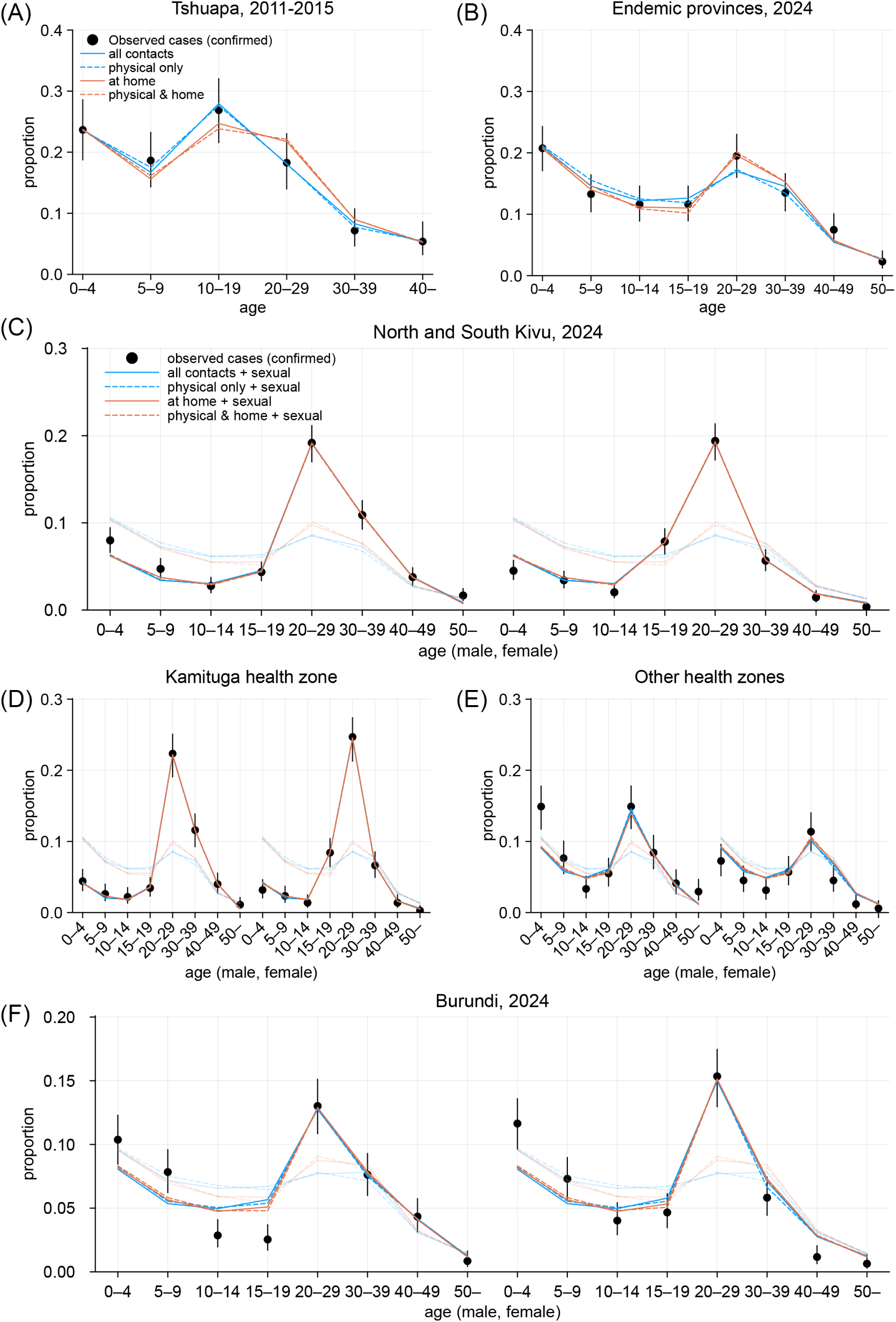
Observed and modelled age distributions of mpox cases. (A) Clade Ia cases with suspected human-to-human exposure from the Tshuapa province, DRC, 2011–2015. Those aged 34 and older were assumed to be smallpox-immunised. (B) Cases from clade Ia-endemic provinces in DRC, January–mid August 2024. Those aged 45 and older were assumed to be immunised. (C)–(F) Clade Ib cases from multiple settings in DRC, January–mid August 2024 and Burundi, mid September–October 2024. Those aged 45 and older were assumed to be immunised. Half-transparent lines represent the model fit without assuming transmission over sexual contact. Dots and whiskers represent the observed age distributions among cases and their 95% confidence intervals.

**Table 1.** Relative contribution of sexual contacts to the transmissibility of clade Ib in DRC and Burundi.

| Model | Contact type | Model weights* | Estimated relative susceptibility** | | Estimated fraction of $R_{\text{eff}}$ attributable to sexual transmission*** | | | |
| --- | --- | --- | --- | --- | --- | --- | --- | --- |
|  |  |  | Children aged 0–4 | Immunised cohort | North and South Kivu | Kamituga health zone | Other health zones, South Kivu | Burundi |
| 1 | All contacts† | 0.21 | 1.9<br>(1.6–2.2) | 0.27<br>(0.18–0.38) | 41%<br>(36–45%) | 60%<br>(54–66%) | 14%<br>(9–21%) | 16%<br>(12–20%) |
| 2 | Physical contacts only | 0.33 | 1.6<br>(1.4–1.9) | 0.30<br>(0.21–0.42) | 42%<br>(37–46%) | 61%<br>(55–66%) | 15%<br>(9–21%) | 16%<br>(13–20%) |
| 3 | All contacts at home | 0.29 | 1.3<br>(1.1–1.6) | 0.22<br>(0.15–0.32) | 43%<br>(38–47%) | 63%<br>(57–69%) | 12%<br>(5–19%) | 15%<br>(11–20%) |
| 4 | Physical contacts at home | 0.17 | 1.2<br>(0.96–1.4) | 0.25<br>(0.17–0.36) | 42%<br>(38–47%) | 63%<br>(57–68%) | 12%<br>(4–19%) | 15%<br>(11–19%) |
| Model average | — | — | 1.5<br>(1.05–2.1) | 0.26<br>(0.17–0.38) | 42%<br>(37–47%) | 62%<br>(55–68%) | 14%<br>(6–21%) | 16%<br>(11–20%) |

Posterior median estimates and 95% credible intervals are shown.
\* Based on the Watanabe-Akaike weights<sup>36</sup>
\*\* Referenced to pre-immunisation cohort aged 5 or older
\*\*\* Defined as the relative reduction in $R_{\text{eff}}$ when sexual contacts were excluded from the next generation matrix
† Physical + conversational contacts. Physical contacts are defined as any skin-to-skin contact and conversational contacts are defined as in-person contacts with a two-way exchange of at least three words.

### Modelling sexual contact transmission for clade Ib outbreaks

Assuming that the community contact transmission patterns relevant to clade Ia would also apply to clade Ib transmission, we modelled a sexual contact transmission route among heterosexual individuals with high sexual activity (who we assume primarily represent FSWs and clients) alongside the community contact matrix to represent the transmission dynamics of clade Ib outbreaks. We considered simple proportionate mixing between age groups, where sexual partners are chosen randomly among those in the high-activity group of the opposite sex. We parameterised the age-dependent proportion of male and female high-activity individuals (aged 15–19, 20–29, 30–39 and 40–49) and their mean neighbour degrees^37^ over the infectious period of mpox to construct the age-, sex- and transmission-route (community vs sexual contact)-dependent next generation matrix. We fitted our model to the age-sex distribution of clade Ib cases in multiple geographical settings: the North and South Kivu provinces (referred to as the Kivus hereafter), the Kamituga health zone, other health zones of South Kivu, and Burundi. Across these geographical settings, we assumed that age-dependent relative susceptibility was identical to what we estimated from clade Ia datasets.

Our model reproduced the observed age-sex distributions of clade Ib cases generally well, including age groups not assumed to be involved in high-activity sexual contact (Figure 1C– F). This supports the hypothesis that the transmission dynamics of clade Ib could be explained by a combination of community contact transmission patterns similar to clade Ia and additional sexual contact transmission among the high-activity group. The estimated role of transmission over sexual contact varied between geographical settings, potentially reflecting different sexual behaviours among high-activity individuals. The estimated relative contribution of sexual contact to the overall effective reproduction number was higher in the Kivus (model averaging estimate: 42% [95% credible interval: 37-47%]), which is most likely attributable to the dynamics in Kamituga health zone (62% [55-68%]) (Table 1). In the other health zones of South Kivu (14% [6-21%]) and Burundi (16% [11-20%]), sexual contact was suggested to play a smaller (although non-negligible) role. A recent study on the Kamituga outbreak associated clade Ib spread in the region with a high number of bars serving as places for commercial sex services in the context of the local gold mining industry^38^. Frequent engagement in commercial sex associated with mining work^39^ in Kamituga and potentially other nearby mining areas^38^ may account for the distinct epidemiological characteristics of clade Ib in the Kivus. These differences in the relative roles between community and sexual contacts could shape contrasting age-dependent transmission patterns between regions. We reconstructed the frequency of transmission across age groups from our modelled next generation matrix assuming proportionate mixing (Figure 2A).

**Figure 2.**
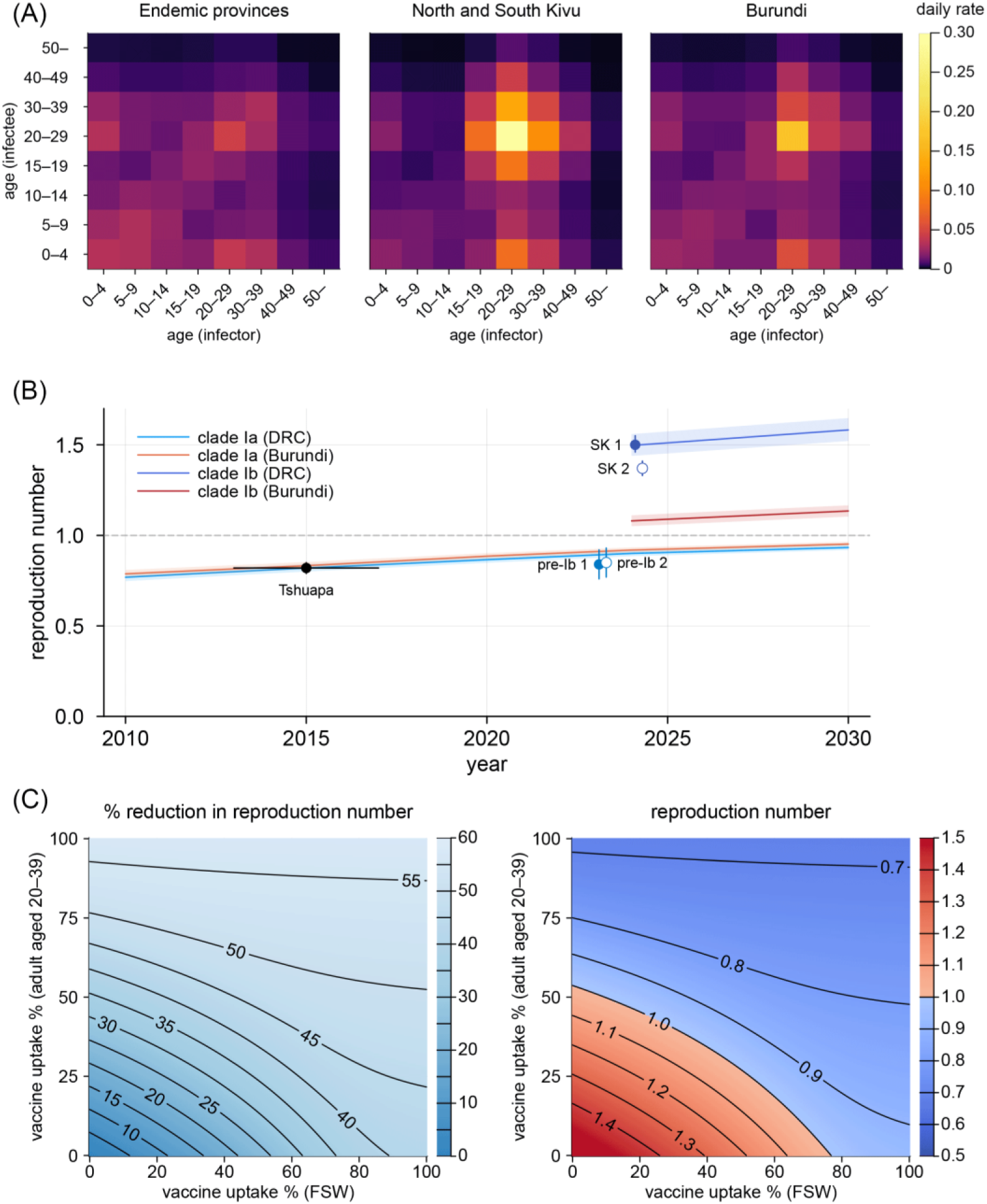
Transmission dynamics of MPXV clade I outbreaks. (A) Age-specific transmission patterns inferred for clade Ia in endemic provinces of DRC and clade Ib in North and South Kivu, and Burundi. Colours represent daily transmission rates between infector-infectee pairs in given age groups according to the estimated next generation matrices. (B) Projected effective reproduction number (*R_y_*) for clades Ia and Ib in DRC (blue lines) and Burundi (red lines), 2010–2030. Lines and shaded areas represent median estimates and 95% credible intervals of *R_y_*. The estimated reproduction number of 0.82 from clade Ia case data between 2013-2017 in Tshuapa province, DRC^21^ was used as a reference value for 2015 (the midpoint of the study period). The empirical estimates of the effective reproduction number from the DRC incidence data in the pre-clade Ib period in 2023 (pre-Ib) and from the South Kivu incidence data (SK) in 2024, as proxy data for clade Ia and Ib, respectively, are displayed as dots for comparison. Paired dots (jittered horizontally for visual aid) denote different serial interval distributions used, which were estimated elsewhere from two separate sets of infector-infectee pairs (distribution 1: mean 17.5 days; distribution 2: mean 11.4 days)^63^. (C) Contour plots for estimated impact of different combinations of FSW-focused and mass vaccination strategies on the effective reproduction number in the Kivus.

Compared with clade Ia in the endemic provinces, clade Ib in the Kivus was estimated to be more frequently transmitted among sexually-active age groups (Figure 2A, left vs middle). Such excess in estimated transmission among the sexually-active age groups was also present but less pronounced in Burundi (Figure 2A, right).

### Linking transmission patterns and outbreak potential of clade I

Our models allowed us to project the time-evolving effective reproduction numbers (*R_y_*) of MPXV clade Ia and Ib (Figure 2B). We constructed *R_y_* to reflect the transmission potential in a hypothetical ‘pre-outbreak’ state, i.e. accounting for immunity from the historical smallpox immunisation but not from infections or mpox vaccination, to focus on the effect of changing smallpox immunity landscape associated with demographic shift. The projection suggested a 9.8% increase in *R_y_* of clade Ia over the last decade in DRC, reflecting the ageing of the smallpox-immunised cohorts. When assuming an *R_y_* of 0.82 for clade Ia in DRC, 2015^21^, this increase translates into an *R_y_* of 0.90 in 2024, which aligns with our estimates from the pre-Ib mpox incidence data in DRC (Figure S2). As *R_y_* gradually approaches 1, the expected final size of an outbreak initiated by a single zoonotic infection, given as 1/(1-*R_y_*)^40^, would have increased from 5.6 in 2015 to 10.0 in 2024, which may also explain the recent rise in clade Ia cases in the endemic areas in DRC^41^.

If the same community contact transmission patterns estimated for clade Ia also apply to clade Ib, the overall *R_y_* in the Kivus combining both community and sexual contact routes would be around 1.5 in our projection. This is similar to our estimate of the initial reproduction number from suspected clade Ib cases in South Kivu (Figure S2). The increase in *R_y_* attributable to sexual contact, resulting in an overshoot of the epidemic threshold of 1, could explain the stable exponential growth of clade Ib cases observed in the Kivus over the summer of 2024. In Burundi, our model projected that *R_y_* for clade Ia would be 4% higher than DRC in 2024 reflecting a slightly different population age distribution. With an additional sexual contact transmission route, *R_y_* for clade Ib in Burundi may have also exceeded the epidemic threshold of 1 (with a point estimate of 1.08), which explains the major spread of clade Ib in Burundi, observed the earliest among the countries neighbouring DRC.

### Focused/mass vaccination strategies

Using the Kivus as a case study, we compared the possible impact of FSW-focused and mass vaccination strategies (Figure 2C). Given the age eligibility for vaccines (18 years and above for JYNNEOS^42^) and the previously smallpox-immunised cohort, we considered adults aged 20–39 as the primary group for mass vaccination—our results also suggested that this group is more likely to contribute to onward transmission in the Kivus (Figure 2A). Under current global vaccine shortages, prioritising FSWs, typically accounting for < 1% of the total population^43^, may allow for the efficient use of available supply. In the Kivus, where sexual contact was estimated to play a substantial role in the overall dynamics, achieving high vaccine uptake among FSWs would reduce the doses required among general adults for control (Figure 2C). Realistically, both strategies may be pursued in tandem, considering uncertainty and a possible shift in the relative role of sexual and community contact transmission, as well as challenges in approaching FSWs facing social barriers and stigma^44^. Regardless of the vaccine target population, clear and supportive communication and de-stigmatisation are essential^45^. Existing community support networks for FSWs and other populations at risk, including HIV/AIDS programmes, could play a key role in public health outreach^45,46^.

## Conclusions

This study quantified the role of community and sexual contacts in MPXV clade I outbreaks. The results suggest that past and current clade Ia outbreaks are primarily driven by community contacts, whereas clade Ib is additionally transmitted through contacts among individuals with high sexual activity such as FSWs and clients. This additional transmission route may be responsible for the increased effective reproduction number of clade Ib in the Kivus compared with clade Ia in other endemic regions. The estimated relative contribution of sexual contact was particularly pronounced in the Kamituga health zone and less so elsewhere, suggesting that the current epidemiology of clade Ib in the Kivus may be largely attributable to sexual contact patterns specific to Kamituga and that generalisability to other settings should be explored in conjunction with the local sociobehavioural contexts. Our results also suggest that, with the ageing of smallpox-immunised cohorts over decades, the reproduction number of MPXV clade I has already been approaching the epidemic threshold of 1 even without additional involvement of sexual contact. In this regard, a small increment in transmission pathways, including but not limited to sexual contact, may sustain a clade I outbreak, particularly in countries with lower historical vaccine coverage.

The current MPXV clade Ib outbreak has exhibited a distinct epidemiological pattern from past clade Ia outbreaks, characterised as more rapid spread involving a novel mode of human-to-human transmission through sexual contact. A similar sudden emergence of sustained human-to-human transmission was the nature of the global clade IIb outbreak in 2022 onwards, which was primarily (if not exclusively) transmitted through sexual contacts among MSM^47^. Our previous modelling work offered a plausible explanation for this transition that clade IIb had established itself in a densely-connected portion of the MSM sexual network, allowing for the rapid spread that was not sustained elsewhere^26^. Likewise, the emergence of clade Ib may be another example of MPXV’s entry into a key population group where it can sustain itself. Whilst extensive transmission among FSWs and their clients was not specifically reported in the clade IIb global outbreak, sexual transmission of clade II involving FSWs had already been reported from 2017^48^ and the potential risk has been discussed since the initial phase of the global outbreak^49,50^. It remains unclear whether only clade Ib is able to be readily transmitted within FSW-related networks or it was just the first to have an opportunity among the clades with similar potential. Recent studies on co-circulating clades Ia and Ib in Kinshasa, DRC^51^ and on the clade IIb outbreak in Sierra Leone^52^ both report overrepresentation of sexually active age groups of both sexes among cases, as was typically observed in clade Ib outbreaks, although without direct evidence of transmission through sexual contacts. Further investigation is warranted to assess and monitor the potential shift in the role of sexual contacts in broader mpox outbreak settings. Regardless, investigation and control efforts for mpox continue to require a tailored and inclusive approach, with a particular focus on the most vulnerable groups including children, MSM, FSWs and other key populations at sexually transmitted infection (STI) risks^53,54^.

### Limitations & sensitivity analyses

This study holds several limitations. First, we assumed that contact survey data sufficiently represented community contacts relevant to MPXV transmission. Some forms of transmission, including fomite or nosocomial routes, may not be fully captured by existing contact data from population samples. Contact matrices also represent the mean contact rates between age groups and neglect possible heterogeneity beyond this. We however believe our eigenvector approach with hold-out validation (Figures S1C and S1E) helped to ensure robustness. We used the empirical contact data from a different country (Zimbabwe) as a proxy since neither empirical contact data from DRC nor Burundi was available.

Nevertheless, the model using Zimbabwe contact data outperformed the model using a synthetic contact matrix for DRC (Extended Data Table 2 and Figure S1) and our findings remain overall consistent between these models (Figure S6).

Second, our model of the sexual contact route for clade Ib was simplistic due to limited sexual behaviour data, e.g. proportionate mixing with age-invariant contact rates. However, the estimated relative contributions of the community and sexual contact routes, the primary interest of this study, would be relatively robust to these assumptions as long as our model can capture the excess in mixing patterns attributable to sexual contacts (Figure S7). We assumed sexual contacts outside of the high-activity groups were captured by contact survey data and had limited contributions to the difference in dynamics between clades Ia and Ib.

Despite our sensitivity analyses, additional data on local sexual behaviours including the detailed distribution of sexual engagement rates in FSWs and clients could further inform the analysis. We also limited our analysis to heterosexual contacts and did not consider MSM networks. The reported clade Ib case profile with near-even sex balance as of the time of analysis (Table S1) does not suggest significant transmission within the MSM group^55^, and our additional analysis also suggests that infection- or vaccine-derived immunity in this group may lower the risk of sustained clade Ib transmission in populations previously affected by the global clade IIb outbreak (Figure S9). However, this does not exclude the future possibility of clade Ib outbreaks among MSM, e.g. in countries without previous clade IIb circulation (including DRC and Burundi^56^) or after herd immunity is lost^57^.

Third, our model primarily focused on a few epidemiological variables (age, sex and sexual activity) as key population attributes and other potential risk factors including malnutrition^58^, co-infection with other STIs (particularly HIV^59^) and living conditions were not explicitly considered. Although some of these factors may be indirectly accounted for by the modelled components, such as age-dependent susceptibility and sexual or community contact matrices, future studies are required to better characterise the role of those risk factors in mpox outbreaks. Biological factors have not been considered either, including possible viral adaptation that might breach our assumption of the same community transmission patterns between clades Ia and Ib. Although our model suggested that the increase in the estimated reproduction number for clade Ib can be explained without requiring evolutionary changes, this does not necessarily exclude their potential role in the clade Ib dynamics.

Finally, case data used for the analysis may have been subject to uncertainty and biases due to challenges in healthcare access, testing or reporting in an outbreak, especially when coinciding with civil conflict and natural disasters^60,61^. Most importantly, the potential underreporting of cases among FSWs, who are facing criminalisation and stigma^62^ may have resulted in an underestimation of the relative contribution of sexual contacts to the clade Ib dynamics.

## Methods

### Data sources

We used the numbers of confirmed mpox cases by age and sex in both historical and current outbreaks from public sources^8,32,64,65^. We used reported cases from 1 January to 18 August 2024 for the current outbreak in DRC for its stable growth trend and the data availability at the subprovincial level (Kamituga and other health zones of South Kivu) during that period. For Burundi which only reported cases from late July 2024 onwards, we used case data from 16 September to 27 October, roughly corresponding to the second half of the stable growth phase until its peak, to exclude potential influences of reporting bias or initial transients.

Further details of the datasets are available in Extended Data Table 1. Weekly incidence data for DRC and South Kivu were collected from public reports^23,24,66^. Empirical contact survey data from Manicaland, Zimbabwe^30^ was retrieved via {socialmixr} R package^67^. Population age distributions in DRC, Burundi and Manicaland from relevant years were collected from censuses (where available)^68,69^ and the World Population Prospects 2024^70^. We also used synthetic contact matrix data published by Prem et al.^71^.

**Extended Data Table 1.** MPXV clade I outbreak datasets.

| ID | Location | Year | Clade | Exposure type | N | Reference |
| --- | --- | --- | --- | --- | --- | --- |
| 1 | Tshuapa province, DRC | 2011-2015 | Ia | Zoonotic* | 308 | 32 |
| 2 | Tshuapa province, DRC | 2011-2015 | Ia | Human-to-human** | 279 | 32 |
| 3 | Six endemic provinces***, DRC | 2024† | Ia | Not classified | 482 | 64 |
| 4 | North and South Kivu provinces, DRC†† | 2024† | Ib | Not classified | 1,377 | 64 |
| 5 | Kamituga health zone, South Kivu†† | 2024† | Ib | Not classified | 725 | 64 |
| 6 | Other health zones, South Kivu†† | 2024† | Ib | Not classified | 510 | 64 |
| 7 | Burundi | 2024††† | Ib | Not classified | 945 | 8,65 |

\* Defined by reporting only exposure to an animal within 3 weeks before onset
\*\* Defined by reporting only exposure to a human with a rash illness only within 3 weeks before onset
\*\*\* Equateur, South and North Ubangi, Sankuru, Thsopo and Tshuapa
† As of 18 August 2024
†† North and South Kivu provinces data also contains cases in Kamituga and other health zones of South Kivu.
††† 16 September–27 October 2024

### Model structure

We assumed that the observed number of mpox cases by age or by age and sex follows a multinomial distribution with probabilities proportional to the elements of the dominant eigenvector of the next generation matrix.

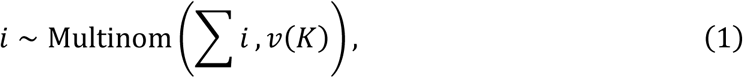

where *v*(*K*) is the normalised dominant eigenvector of the next generation matrix *K*. We constructed the next generation matrix as a product of transmissibility *β*, age-dependent relative susceptibility *σ_a_* and a contact matrix *C*. We used age-disaggregated case data and contact matrices for the analysis of clade Ia; i.e. *C* = {*c<u>_ab_</u>*}, where *c_ab_* represents the mean number of community contacts (conversational and/or physical, depending on the type of contact matrix used) an individual from age group *b* has per unit time with those from age group *a*. We used multiple contact matrices potentially relevant to mpox transmission (Extended Data Table 2). For each contact matrix, *β* was adjusted so that the dominant eigenvector of the next generation matrix estimated for the historical Tshuapa dataset is equal to the previously estimated reproduction number of 0.82^21^.

For the analysis of clade Ib, we used age-and-sex-disaggregated case data and expanded the contact matrix to account for heterosexual transmission dynamics in addition to community contacts. We stratified male and female populations into high and low sexual activity groups. Following the approach in Endo et al.^26^, we further stratified cases in the high-activity groups by their route of exposure (community-vs sexually-associated exposures) and allowed sexually-acquired cases to have higher risks of onward sexual transmission because of a substantial variance in sexual behaviour^72,73^. Namely, in a highly heterogeneous sexual network, transmissions through sexual contact tend to concentrate among individuals with the highest rates of sexual activity; this renders sexual exposure an indicator of even higher contact rates than the average among the high-activity groups. We assumed that this variance effect is negligible among low-activity groups (such that stratification by exposure route can be disregarded) and that their sexual contacts are reflected in the existing contact matrices, whose definition of contacts includes sexual contacts. Despite its simplicity, our approach of stratifying sexual contacts by two activity classes and two exposure routes could well capture the stationary growth pattern of mpox transmission (see Supplementary Information for a proof-of-concept network model).

Let subscripts *H* and *L* denote high- and low-activity groups and *M*/*m* and *F*/*f* represent male and female cases with sexual (upper-case) and community exposures (lower-case), respectively. Vectors *i*_X_ represent the age-stratified number of mpox infections in category *X*. The generation-wise reproduction process of *i*_X_’s is then described in a block matrix format (each block is an 8-by-8 matrix accounting for age group stratification) as:

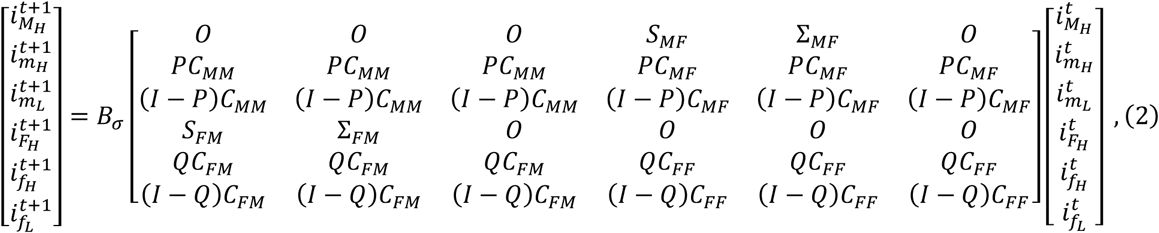

where *O* and *I* are zero and unit matrices, respectively. *S*_MF_ and *S*_FM_ are block contact matrices representing heterosexual transmission from sexually-acquired high activity individuals. Similarly, *Σ*_MF_ and *Σ*_FM_ represent heterosexual transmission from community-acquired high activity individuals. Again, note that we expect *S*_X_ to be larger than *Σ*_X_ because sexually-acquired cases are more likely to have more sexual partners than the population average. *P* = (*p_a_*) and *Q* = (*q_a_*) are diagonal matrices representing the age-dependent proportions of high-activity individuals for males and females, respectively. *C*_X_ is a sex-specific contact matrix and is assumed to be half of the sex-aggregated contact matrix hereafter for simplicity. We define *B_σ_* as a diagonal block matrix whose diagonal blocks are an 8-by-8 diagonal matrix of the age-specific susceptibility *σ_a_* multiplied by transmissibility *β*.

Noting that for *t* ≥ 1, *i*_*mH*_ = *P*(*i*_*mH*_ + *i*_*mL*_) and *i*_*fH*_ = *Q*(*i*_*fH*_ + *i*_*fL*_), the above relationship can be simplified into

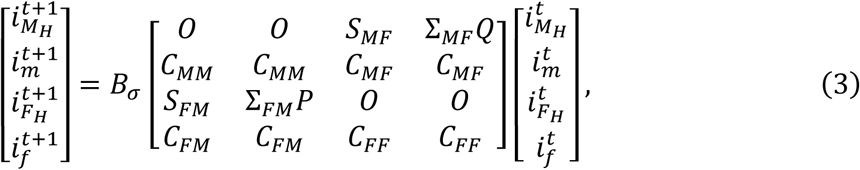

where *i*_*m*_ = *i*_*mH*_ + *i*_*mL*_ and *i*_*f*_ = *i*_*fH*_ + *i*_*fL*_.

We assumed proportionate mixing to model *S*_MF_, *S*_FM_, *Σ*_MF_ and *Σ*_FM_; further details on parameterisation are described in the next section.

We constructed a multinomial likelihood for the observed number of male and female cases by age group:

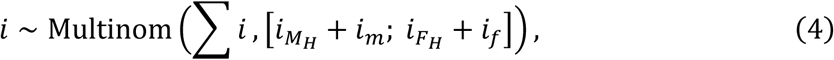

where [*i*_*MH*_ + *i*_*m*_; *i*_*FH*_ + *i*_*f*_] is a vector obtained by sex-wise aggregation of the normalised dominant eigenvector of the next generation matrix in Equation 3.

**Extended Data Table 2.** Community contact matrices used for analysis.

| ID | Contact type | Contact setting | Data type | Country | Reference | Hold-out validation loss* | Hold-out empirical KLD** |
| --- | --- | --- | --- | --- | --- | --- | --- |
| 1 | All | All | Empirical | Zimbabwe | 30 | 47.23 | 0.009 |
| 2 | Physical only | All | Empirical | Zimbabwe | 30 | 47.18 | 0.009 |
| 3 | All | Home | Empirical | Zimbabwe | 30 | 44.37 | 0.006 |
| 4 | Physical only | Home | Empirical | Zimbabwe | 30 | 46.40 | 0.008 |
| 5 | All | All | Synthetic | DRC | 71 | 79.60 | 0.04 |
| 6 | All | Home | Synthetic | DRC | 71 | 62.68 | 0.03 |
| 7 | All | Home | Synthetic | Burundi | 71 | NA*** | NA*** |
| 8 | All | All | Synthetic | Burundi | 71 | NA*** | NA*** |

\* Mean negative log-likelihood times two for the validation dataset (Dataset 3, Extended Data Table 1) not used in training
\*\* Kullback-Leibler divergence<sup>74</sup> from the empirical distribution of the validation dataset to the mean modelled distribution
\*\*\* Not used for validation; only for the sensitivity analysis (Figure S6)

### Model fitting

Our model fitting process was two-fold. We first fitted and validated the model of community contact transmission using the age distributions of cases from the historical and current clade Ia outbreaks in DRC. We used this model to estimate age-dependent susceptibility parameters *σ_a_* for the community contact matrix. We then constructed an expanded contact matrix including sexual contacts and calibrated it to the age-sex distributions of cases from clade Ib outbreaks in DRC and Burundi in order to estimate additional parameters that describe transmission dynamics of clade Ib through community and sexual contact routes.

#### (i) Analysis of clade Ia outbreaks

As in the previous section, we defined the next generation matrix *K*={*k_ab_*}=*βσ_a_c_ab_*. The contact matrix *c_ab_* was constructed from existing datasets (Extended Data Table 2), with adjustment for age structures between the source and modelled populations where different (using the density correction method described by Arregui et al.^75^). Based on data availability, we used 6 age groups (0-4, 5-10, 10-19, 20-29, 30-39 and 40+) for the 2011–2015 Tshuapa data and 8 age groups (0-4, 5-10, 10-14, 15-19, 20-29, 30-39, 40-49 and 50+) for the 2024 DRC endemic provinces data. When the age groups in the case data were coarser than the contact matrix, the elements were merged accordingly^76^. Transmissibility *β* is a nuisance scaling parameter and was not estimated. We assumed that children aged 0-4 years may have potentially higher susceptibility to infection and that adults born before the cessation of the smallpox vaccine in 1980^14^ may have lower susceptibility. To account for this, we parameterised *σ_a_* as

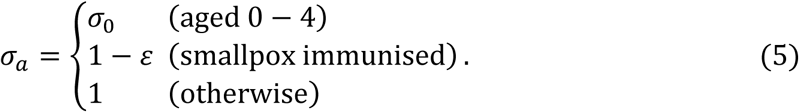

Here, *ε* represents the net vaccine protection among the vaccinated cohorts, i.e. the product of the vaccine effectiveness and coverage in the historical campaign (also referred to as the effective vaccine coverage). The susceptibility for the age group that contains both vaccinated and unvaccinated cohorts (ages 30-39 in the historical Tshuapa dataset and the ages 40-49 in the 2024 outbreak datasets) was specified as the weighted average between 1-ε and 1, based on the proportion of individuals born in or before 1980 in those age groups^70^ as of 2013 (the midpoint of the Tshuapa dataset period) or 2024. Given the high historical vaccination coverage in DRC (89–97%)^35^, the estimated vaccine protection *ε* would provide an approximation (or at least a reasonable lower bound) for the effectiveness against clade I mpox. We estimated *σ*_0_ and *ε* by the Bayesian importance sampling method^77^ based on the multinomial likelihood in Equation 1 and obtained median estimates and 95% credible intervals (CrIs).

The exploratory model development was done using the historical Tshuapa dataset. The resulting model, described above, was then validated using the 2024 DRC endemic provinces dataset. We estimated two parameters *σ*_0_ and *ε* from the historical Tshuapa dataset alone and compared the model outputs (after accounting for a decade’s shift in the age of vaccinated cohort) with the age distribution from the (hold-out) 2024 DRC endemic provinces dataset (Figure S1). The model outputs showed good overall concordance with the validation data. We obtained final parameter estimates from the joint estimation using both datasets for the subsequent analyses.

#### (ii) Analysis of clade Ib outbreaks

We parameterised the block components representing sexual contacts among high-activity individuals *Σ*_MF_, *Σ*_FM_, *S*_MF_ and *S*_FM_ in Equation 3 as follows. We assumed age groups of 15-19, 20-29, 30-39 and 40-49 years potentially contain high-activity populations, i.e. *p_a_* = *q_a_* = 0 outside those age groups. We assumed a simple proportionate mixing assumption (i.e. no age assortativity) where each high-activity individual makes sexual contacts at a specified rate over the infectious period of MPXV, which are randomly assigned to other high-activity individuals of the opposite sex irrespective of age. We denote the mean sexual contact rates among high-activity females and males infected through community contact as *v*_F_ and *v*_M_; similarly, we denote the mean sexual contact rates among those infected through sexual contact as *w*_F_ and *w*_M_. Note that *v*_F_, *v*_M_, *w*_F_ and *w*_M_ are defined to have the same scale as the community contact matrix in relation to onward transmission, i.e. one unit of *v*_F_, *v*_M_, *w*_F_ and *w*_M_ represent the amount of sexual contact that contributes to transmission equivalent of one daily community contact. The entries of *Σ*_MF_, *Σ*_FM_, *S*_MF_ and *S*_FM_ are then modelled as:

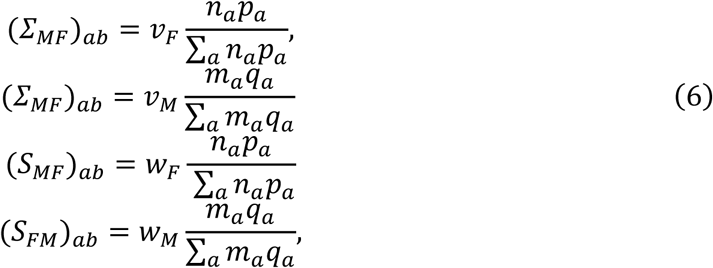

where *n_a_* and *m_a_* are the relative population size for males and females by age group (we assumed even sex distribution^78,79^, i.e. ∑_*a*_ *n*_*a*_ = ∑_*a*_ *m*_*a*_ = 1/2). Note that *S*_X_ and *Σ*_Χ_ are proportional with a ratio of *w*_X_ : *v*_X_. We assumed that infection through community contact has no association with one’s sexual contact rate, i.e. *v*_F_ and *v*_M_ represent the mean sexual contact rates among the high-activity groups. Reciprocity of heterosexual contacts then requires:

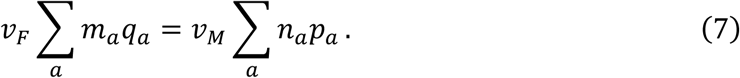

We estimated the ratio between *w*_F_ and *v*_F_ assuming that the sexual network among high-activity groups is represented by a configuration network (i.e. no degree assortativity; an alternative scenario is discussed in the Supplementary Information), in which *v*_X_ corresponds to the mean degree and *w*_X_ to the mean neighbour degree (or degree-weighted average of degree)^37^. We assumed female sex workers (FSWs) represent the behaviour of the majority of high-activity female individuals and used an FSW survey data from DRC^80^. Since both the mean degree and mean neighbour degree can be characterised by the mean *μ* and standard deviation (SD) *σ* of the sexual contact degree distribution *π*(*x*), their ratio is estimated as:

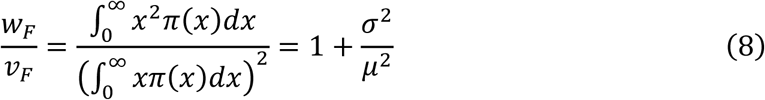

We used the reported mean and SD for the weekly number of clients (mean: 17.61; SD: 12.0) and non-paying partners (mean: 5.51; SD: 22.0) per FSW^80^ to derive *w*_F_/*v*_F_ = 2.17. Here we assumed that the numbers of clients and non-paying partners are uncorrelated (an alternative scenario discussed in Supplementary Information); the sexual contact degree as a sum of these two numbers would then have a mean 17.61 + 5.51=23.12 and a variance of 12.0^2^ + 22.0^2^ = 628, which were supplied to Equation 8. This and the reciprocity requirement in Equation 7 assure that *v*_M_ and *v*_F_ are derived from *w*_F_, {*p_a_*} and {*q_a_*}. We also set boundary conditions for the total number of high-activity individuals, i.e. ∑_*a*_*n*_*a*_*p*_*a*_ and ∑_*a*_ *m*_*a*_*q*_*a*_. Namely, we assumed that high-activity males account for 10% of the male population among the sexually-active age groups and high-activity females 1% of the total female population, based on engagement data in commercial sex in Africa (proportion of reporting paid sex in the last 12 months among males aged 15-59 in Mozambique^81^ and the estimated number of FSWs per capita in DRC^43^).

Assuming the community transmission patterns are similar between clades Ia and Ib, we used the same *σ_a_* estimated for clade Ia throughout the clade Ib analysis. This implicitly assumes uniform smallpox vaccine coverage across geographical settings (see Figure S7 for sensitivity analysis). The rest of the parameters, {*p_a_*}, {*q_a_*}, *w*_F_, *w*_M_ were estimated by the Markov-chain Monte Carlo (MCMC) method (No-U-Turn Sampler^82^). We obtained 2,000 MCMC samples, while ensuring the effective sample sizes (ESS) of at least 500 for every parameter; most parameters achieved an ESS of ∼1,000 or larger. Using these posterior samples, we estimated the proportion of *R*_eff_ attributable to sexual transmission as a measure of relative contribution of sexual contacts to mpox transmission. We defined it as the relative reduction in *R*_eff_ by excluding sexual contacts from the next generation matrix. Namely, this proportion is given as 1 - *λ*_C_/*λ*_SC_, where *λ*_SC_ and *λ*_C_ are the dominant eigenvalue of the next generation matrix (see Equation 3) with and without the sexual contact blocks (*S*_X_ and *Σ*_X_), respectively.

**Extended Data Table 3.**
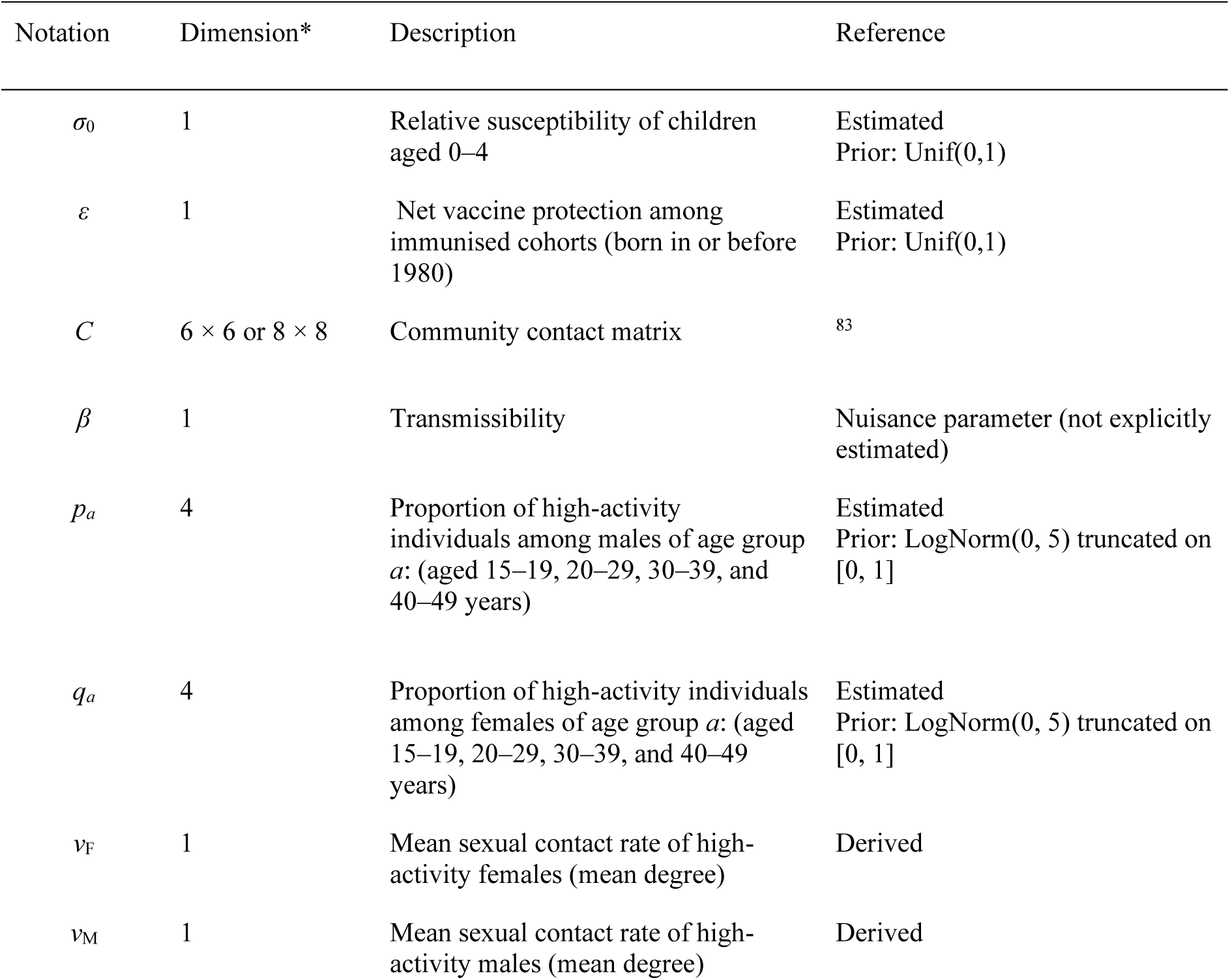
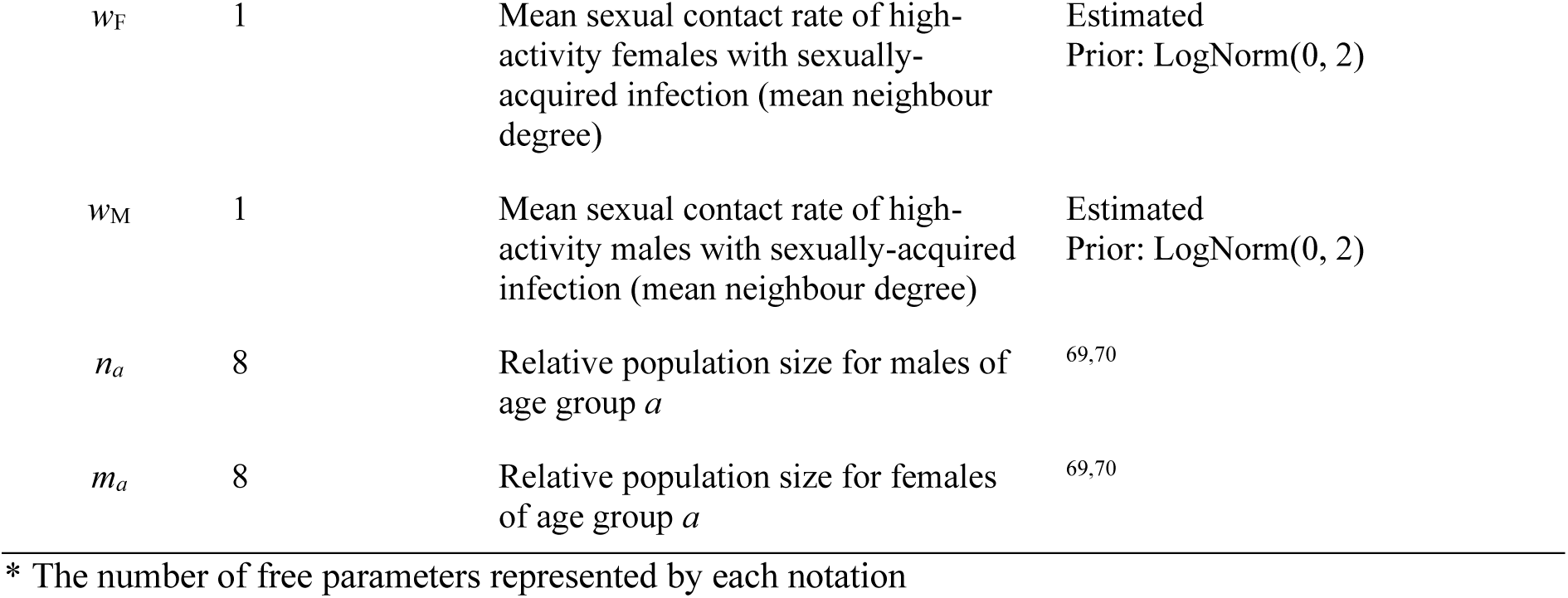
Model parameters.

### Model averaging and outputs

We used model averaging over candidate models using different contact matrix data (Extended Data Table 2) with Watanabe-Akaike weights^36^ based on the combined likelihood for datasets 2, 3 and 4 in Extended Data Table 1. We did not include the likelihood for subprovincial datasets (datasets 5 and 6) or Burundi (dataset 7) in the weights for the main analysis due to overlapping cases and potentially different data collection and contexts, respectively. Alternatively, weights also using these likelihoods were included in our sensitivity analysis (see Supplementary Information). Models 5 and 6 using synthetic contact matrices had substantially worse likelihood than other models resulting in effectively zero weights and thus were not included in the main analysis.

#### (i) Frequency of transmission between age groups

We estimated the relative frequency of infector-infectee age pairs at equilibrium using the next generation matrix and the normalised dominant eigenvector:

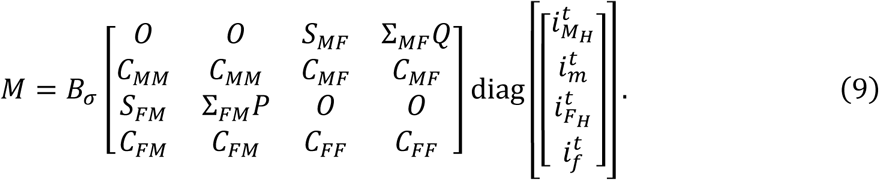

We then aggregated the block elements of *M* to obtain a sex- and transmission-route aggregated frequency matrix.

#### (ii) Projection of time evolution of the reproduction numbers

We projected the effective reproduction number (*R_y_*) of mpox in DRC and Burundi by shifting the age of the smallpox-immunised cohorts and modifying the contact matrices^75^ according to the population projection^70^. Referring to the previous estimate^21^, we assumed the effective reproduction number of clade Ia in the Tshuapa province was 0.82 in 2015 (midpoint of the estimation period 2013–2017). We then projected the effective reproduction number for clade Ia between 2010 and 2030 as the dominant eigenvalue of the next generation matrix for each year, accounting for the following two factors. First, the relative susceptibility of age groups that include the immunised cohorts was adjusted according to the estimated net protection *ε* (model averaging weights from Table 1) and the proportion of immunised cohorts (those born in or before 1980 as per the population projection^70^) within the age groups. Second, the community contact matrices are also adjusted to the projected population age distribution by the density correction method^75^. For clade Ib, we projected the effective reproduction number in 2024 onward assuming the patterns of community contact transmission are shared between clades Ia and Ib. Namely, the *R_y_* for clade Ib was projected by incorporating additional sexual contact contributions described in Equation 3 (estimated for the Kivus and Burundi) into the projected next generation matrices for clade Ia, assuming time-invariant sexual contact patterns. Here we assumed a hypothetical ‘pre-outbreak’ state and neglected potential infection- or mpox vaccination-derived immunity as well as any future interventions.

#### (iii) The effective reproduction numbers under focused and mass vaccination scenarios

We modelled the change in the effective reproduction number of clade Ib under a combination of FSW-focused and mass vaccination scenarios. We used an mpox vaccine effectiveness estimate of 86%^84^, which we assumed to protect against infection but not against onward transmission in the case of breakthrough infections. FSW-focused vaccination was assumed to be offered to female high-activity individuals of age 20–39. For mass vaccination, all individuals aged 20–39 years, regardless of sexual activity levels, were included. The baseline effective reproduction numbers without vaccines were matched to the values projected in (ii) for each population setting.

## Data Availability

All data used in this study are publicly available. All the analysis was conducted in Julia v.1.9.3 and R v.4.3.1. Source codes are available on a GitHub repository: https://github.com/akira-endo/mpoxclade1_eigen.

## Acknowledgement

H.M., T.R.A., S-m.J. and A.E. are supported by the Japan Science and Technology Agency (JST) (JPMJPR22R3, to A.E.) and Japan Agency for Medical Research and Development (JP223fa627004). H.M. and A.E. are supported by the Japan Society for the Promotion of Science (JSPS) (JP22K17329, to A.E.). H.M., J.H.F. and A.E. are supported by Cabinet Agency for Infectious Disease Crisis Management, Cabinet Secretariat of Japan. T.R.A. is also supported by the Rotary Foundation (GG2350294), the Nagasaki University World-leading Innovative & Smart Education (WISE) Program of the Japanese Ministry of Education, Culture, Sports, Science and Technology (MEXT) and the JSPS (JP24KJ1827). KE is supported by the Ministry of Education, Singapore, under the Academic Research Fund Tier 1 Seed Award (RLMOE100201900000001), a Singapore Ministry of Education startup grant (LKCMedicine-SUG, #022487-00001) and JST (JPMJPR23R3). S-m.J. is supported by the Centers for Disease Control and Prevention (CDC) Safety and Healthcare Epidemiology Prevention Research Development programme (200-2016-91781). A.E. is also supported by National University of Singapore Start-Up Grant. A.N. and A.E. are supported by the National Institutes of Health (K01AI166347, to A.N.), the National Science Foundation (NSF #2230125, to A.N.). A.N. is also supported by JST (JPMJPR21R8). S.J. and B.L.D. are supported by the Ministry of Education Reimagine Research Grant and Programme for Research in Epidemic Preparedness and REsponse (PREPARE), Ministry of Health, Singapore. J.H.F is supported by PREPARE.

## Author contribution

Conceptualisation: A.E. Methodology: H.M., T.R.A, K.P. and A.E. Investigation: H.M., T.R.A and A.E. Visualisation: H.M., T.R.A and A.E. Data curation: T.R.A., S.J., K.P. and A.E. Funding acquisition: A.E. and A.N. Writing – original draft: H.M., T.R.A and A.E. Writing – review and editing: B.L.D., D.B., J.H.F., S.J., P.K.M., K.E., S-m.J., A.N. K.P., A.M.W., D.S.N. and D.N.

## Footnotes

a Contact involving a minimum threshold of words exchanged during a two-way conversation without any physical contact

