## Supplementary Information for "Roles of community and sexual contacts as drivers of clade I mpox outbreaks"

### Supplementary text: Additional results

#### *Model validation results*

A null model that assumed constant age-dependent susceptibility (i.e. the age distribution of cases should follow the eigenvector of the original contact matrices) did not produce a good fit to the historical Tshuapa dataset particularly in age groups 0–4 and 30+ (Figures S1A and B). A model with two parameters $\sigma_0$ and $\varepsilon$ (susceptibility for age 0–4 and vaccine protection) trained on the Tshuapa dataset showed an improved fit (Figures S1C and D). The model validation using the 2024 endemic provinces dataset, which was not used in model development and training, was overall supportive of our models with a preference for the use of empirical contact matrices over synthetic contact matrices (Extended Data Table 2), where a difference of 10 or more suggests strong support[1].

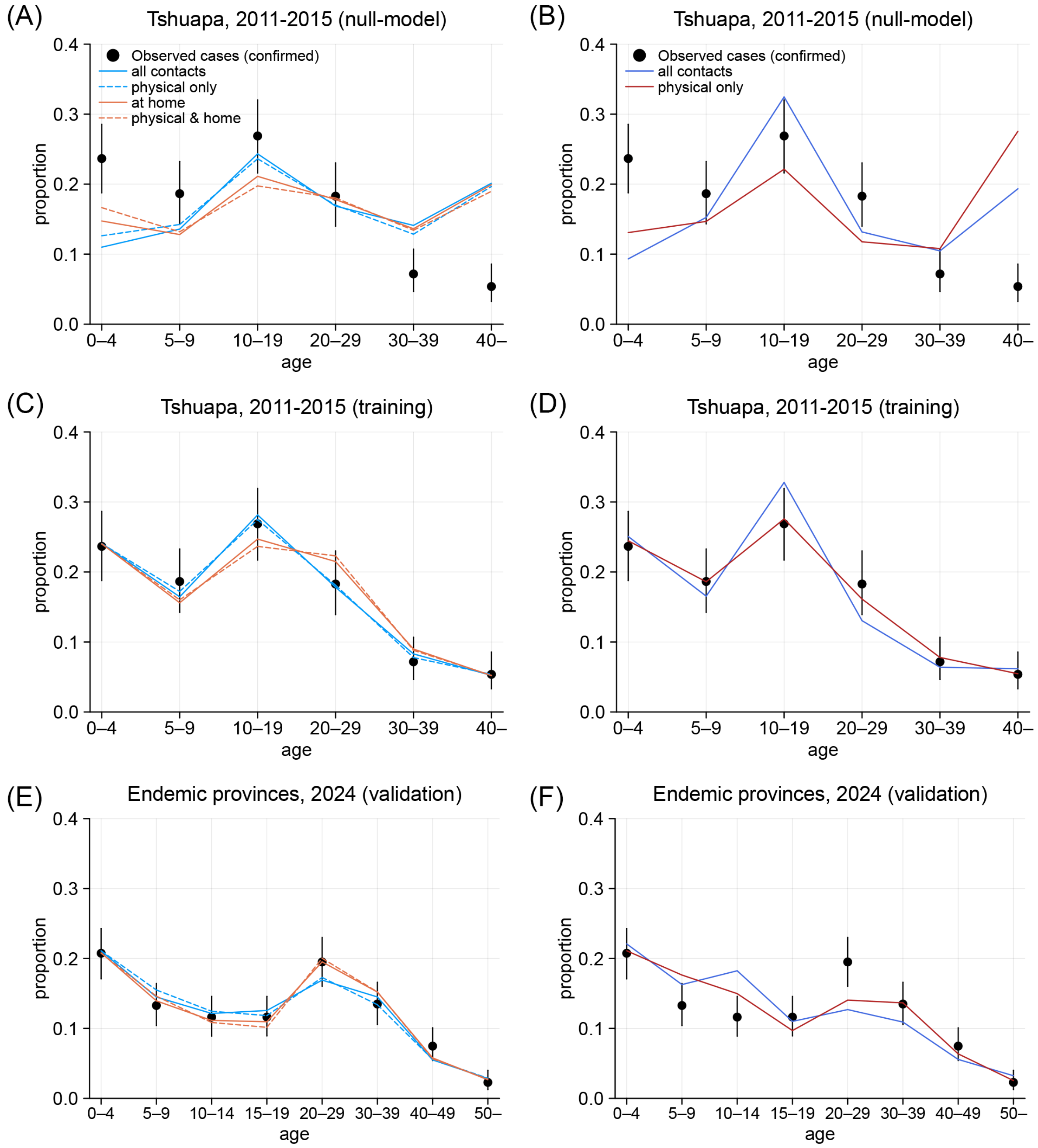

**Figure S1. Model training and validation.** (A) Null-model outputs for the historical Tshuapa dataset, 2011–15 using empirical contact matrices and (B) using synthetic contact matrices. (C) Model training outputs for the historical Tshuapa dataset, 2011–15 using empirical contact matrices and (D) using synthetic contact matrices. (E) Model validation outputs for the endemic provinces dataset, 2024, using empirical contact matrices and (F) using synthetic contact matrices.

##### *Effective reproduction numbers for clade Ia and Ib in DRC from the recent data*

We estimated the effective reproduction number ($R_{\mathrm{eff}}$) for clades Ia and Ib from the recent incidence data with the renewal equation following Marziano et al.[2]. The reported incidence from DRC in September 2023 onward contains both clades Ia and Ib. We therefore estimated $R_{\mathrm{eff}}$ for clade Ia by using only the confirmed number of cases in DRC between January to August 2023[3]. We assumed a constant zoonotic spillover rate of 3.07 cases per week for clade Ia in DRC based on the previously-estimated rate of reported spillovers in the Tshuapa province, DRC[4]. Namely, we translated the original spillover rate of 33 cases per year, accounting for different scales of outbreaks between the Tshuapa data and the DRC data during the pre-clade Ib period of interest (Figure S2B) in terms of average incidence rates (934 cases over 5 years vs 601 cases over 8 months). We multiple-imputed weekly spillover cases using a Poisson distribution with a mean of 3.07 truncated at the observed case counts. We repeated the imputation 100 times to obtain a pooled estimate of $R_{\mathrm{eff}}$. For $R_{\mathrm{eff}}$ of clade Ib, we used suspected case data from South Kivu reported in the DRC situation reports[5,6]. We assumed no zoonotic spillover in South Kivu, where mpox was not endemic before the emergence of clade Ib. We used two serial intervals, each estimated as a Weibull distribution from a separate set of infector-infectee pairs in Marziano et al.[2].

Our $R_{\mathrm{eff}}$ estimates for clade Ia between January to August 2023 in DRC were 0.84 (0.76–0.92) and 0.85 (0.77–0.93) using estimated serial interval distributions: distribution 1 with a mean of 17.5 days (from two household outbreaks in Sudan in 2005 and in Central African Republic in 2021-2022) and distribution 2 with a mean of 11.4 days (from a hospital-associated outbreak in the Republic of the Congo in 2003) in Marziano et al.[2], respectively (Figure S2D). Median estimates of $R_{\mathrm{eff}}$ for clade Ib in 2024 in South Kivu were 1.50 (1.46 - 1.55) and 1.37 (1.33 - 1.41) for each serial interval, respectively.

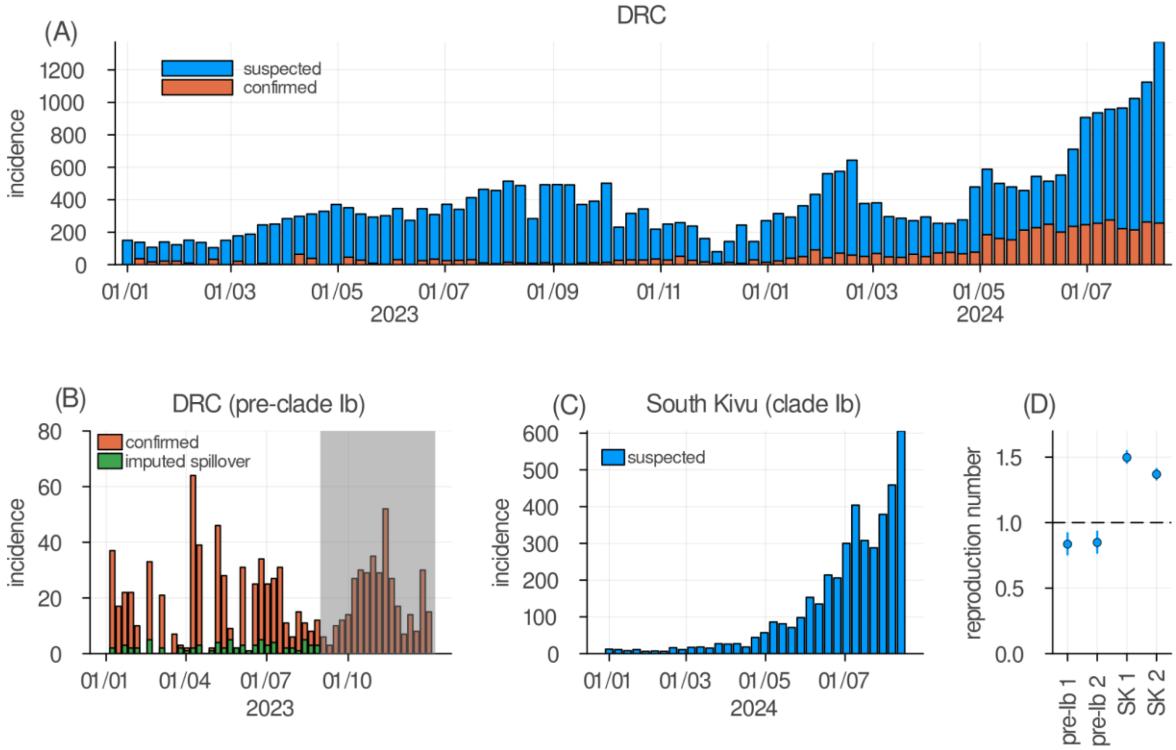


**Figure S2. Epidemic curves and effective reproduction numbers for clades Ia and Ib in DRC.** (A) Epidemic curves for suspected and confirmed cases in DRC in 2023–2024. (B) Epidemic curve of confirmed cases along with imputed zoonotic spillover cases in DRC in 2023. Incidence in the pre-clade Ib period (before September 2023) was assumed to represent clade Ia. Data in the shaded period was excluded from $R_{\mathrm{eff}}$ estimation due to combined reporting of clades Ia and Ib. Green bars shown are one realisation of the imputations. (C) Epidemic curve of suspected cases in South Kivu, 2024, assumed to represent clade Ib. (D) Estimated $R_{\mathrm{eff}}$ for the clade Ia and Ib using pre-clade Ib DRC and South Kivu incidence data, respectively. We used different serial interval distributions estimated from two separate sets of infector-infectee pairs for estimation[2], denoted by suffixes 1 and 2.

***Estimated impact of FSW-focused and mass vaccination strategies in Burundi on the effective reproduction number***

Following the approach of Figure 2C in the main text, we also projected the possible impact of different combinations of FSW-focused and mass vaccination strategies on effective reproduction number in Burundi. Caution is warranted given the uncertainty due to limited sample size and parameter correlation for Burundi (Figure S5).

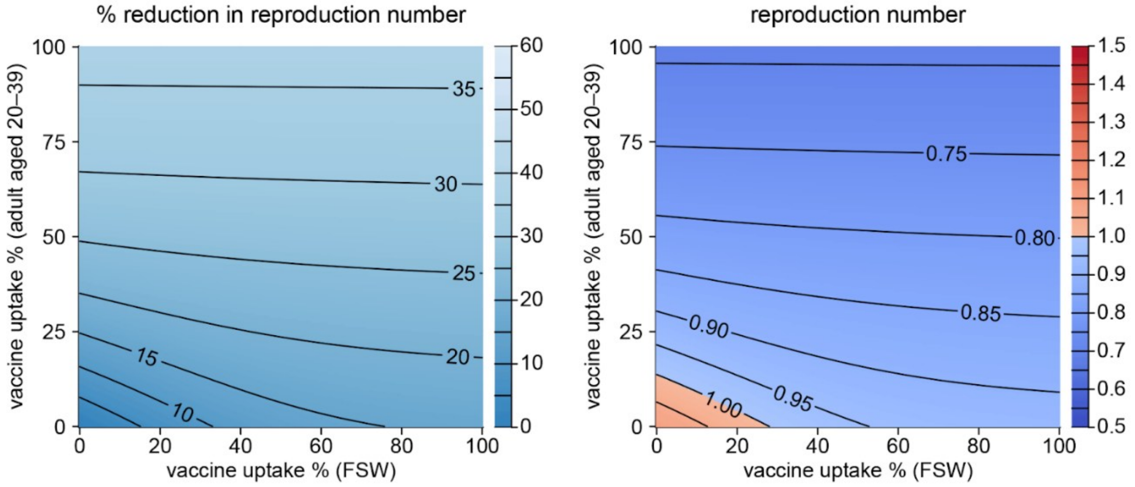


**Figure S3. Estimated vaccine impact in Burundi.** Comparison of the estimated impact between focused vaccination for female sex workers (FSWs) and mass vaccination strategies on the effective reproduction numbers in Burundi.

##### *Modelled sexual contact patterns*

Using posterior samples from our model fitting, we visualised several aspects of inferred sexual contact patterns in clade Ib outbreaks. Caveats must be noted given uncertainties in model assumptions on sexual contact patterns (e.g. proportionate mixing assumption) and limited data; these results are presented to provide contexts in our model and should not be viewed as conclusive evidence on specific sexual behaviours in the populations analysed.

Figure S4 shows the inferred distribution of age groups among high-activity males and females. Although large uncertainties exist, particularly for other health zones of South Kivu and Burundi with smaller estimated sexual contact contributions, the age distributions suggested an overall similarity across locations. Males of age 20–49 and females of age 15–29 account for a large proportion of high-activity groups, which aligns with typical age profiles associated with higher STI risks[7,8].

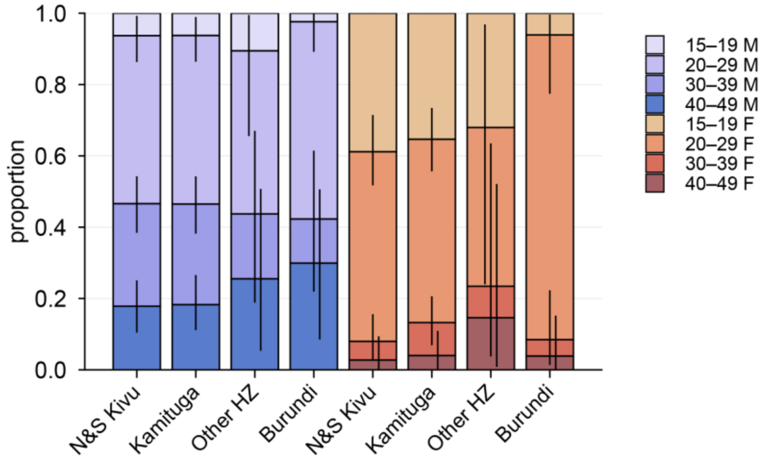


**Figure S4. Estimated age distributions within high-activity male and female groups.** Bars and whiskers represent the posterior median and 95% credible intervals for the North and South Kivu provinces (N&S Kivu), Kamituga health zone of South Kivu, other health zones (Other HZ) of South Kivu and Burundi.

We overall observe near-even sex balance among clade Ib cases across locations (Table S1), both cases of all ages and of sexually active ages (15–49 years). The mean neighbour degrees ($w_F$ and $w_M$) primarily characterise the sexual contact transmission in our model. Compared with $w_F$, the estimates for $w_F$ had more non-overlapping credible intervals (CrIs) between locations, suggesting that the different seeking behaviour for sexual contacts among males may have contributed more to the different relative role of sexual transmission in the clade Ib dynamics. We also report the geometric mean and the ratio for $w_F$ and $w_M$ as their posterior correlation plots suggested non-identifiability for the datasets from other health zones of South Kivu and Burundi (Figure S5). These shapes of the posterior distributions indicated that methods assuming asymptotic normality cannot be used for model selection/averaging. We therefore used Watanabe-Akaike information criterion (WAIC)[9] throughout our study, which does not rely on the normality assumption. The WAIC values strongly supported inclusion of additional sexual contacts with a large difference of 10 to 500 across the modelled locations (Table S1).

**Table S1. Estimated sexual contact patterns**

| | | North and South Kivu | Kamituga health zone | Other health zones, South Kivu | Burundi |
|---|---|---|---|---|---|
| Cases by sex (male/female) | All age | 762 (55%) / 615 (45%) | 375 (52%) / 350 (48%) | 315 (62%) / 195 (38%) | 467 (49%) / 478 (51%) |
| | Age 15–49 | 526 (53%) / 473 (47%) | 300 (50%) / 298 (50%) | 168 (59%) / 116 (41%) | 260 (50%) / 255 (50%) |
| Model-averaged mean neighbour degree (normalised*) | Male ($w_M$ / $\lambda_C$) | 1.4 (1.1–1.8) | 2.6 (2.0–3.4) | 0.2 (0.02–0.9) | 1.2 (0.7–1.9) |
| | Female ($w_F$ / $\lambda_C$) | 2.1 (1.7–2.6) | 2.7 (2.1–3.2) | 3.3 (1.3–5.8) | 1.2 (0.7–2.1) |
| | Geometric mean ($\sqrt{w_M w_F}$ / $\lambda_C$) | 1.7 (1.6–1.9) | 2.6 (2.3–3.2) | 0.9 (0.3–1.2) | 1.2 (1.1–1.3) |
| | Ratio ($w_M$ / $w_F$) | 0.8 (0.7–1.02) | 0.98 (0.8–1.2) | 0.3 (0.1–0.8) | 1.02 (0.6–1.7) |
| ΔWAIC for models without sexual contacts**, by contact matrix types | All contacts | 136.8 | 495.9 | 28.8 | 99.0 |
| | Physical contacts only | 131.4 | 503.7 | 30.0 | 101.7 |
| | All contacts at home | 89.1 | 415.7 | 13.0 | 63.6 |
| | Physical contacts at home | 76.4 | 406.0 | 10.5 | 56.8 |

Ranges show 95% credible intervals.
* Relative to the dominant eigenvalue of the community contact matrix ($\lambda_C$) to account for different scales between contact matrix types (all contact / physical only / home contact / physical & home)
** Deterioration in Watanabe-Akaike information criterion when community-contact only models were used instead of the main models with both sexual and community contacts. Large positive values indicate that WAIC strongly supported the main models.

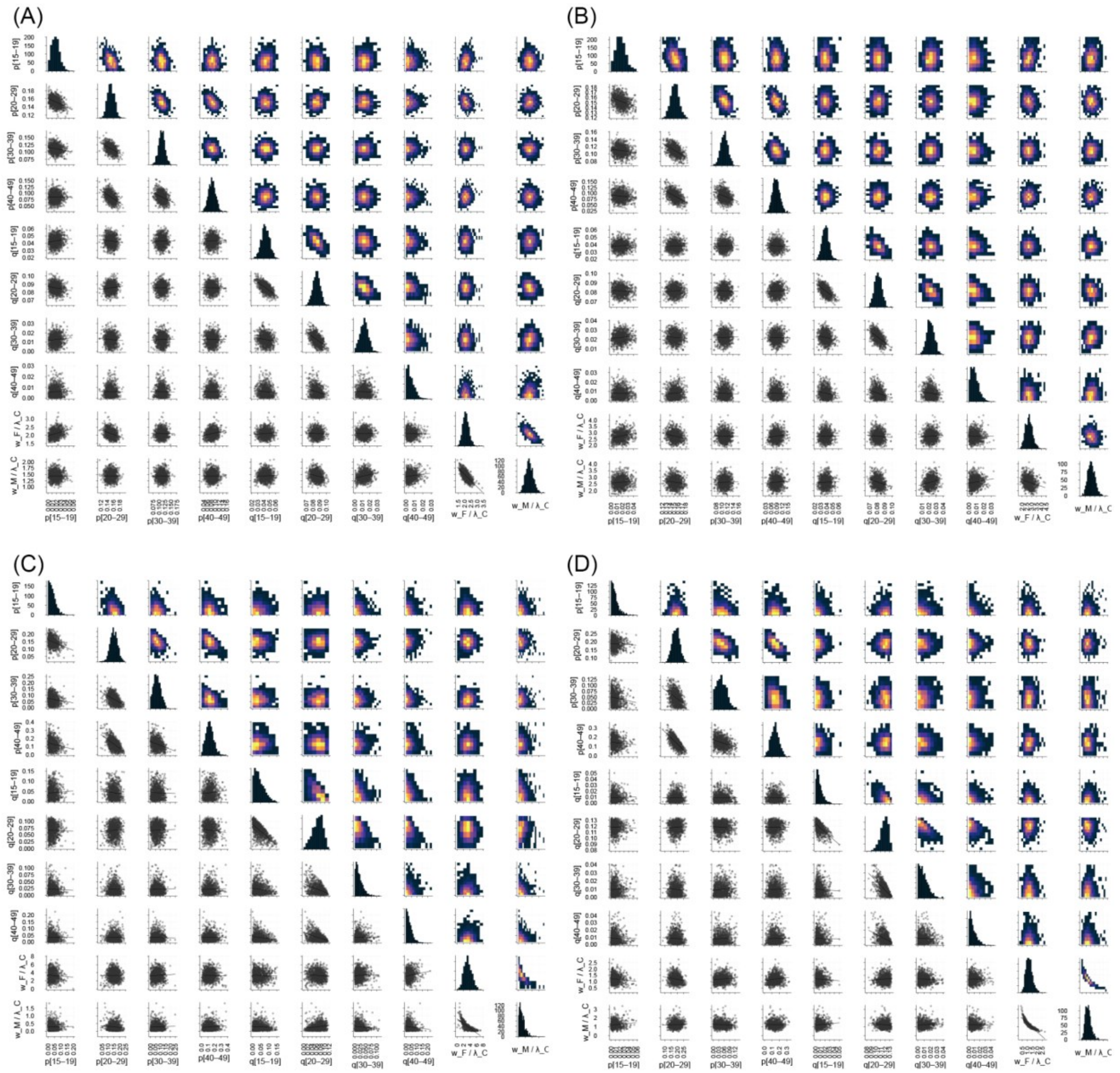


**Figure S5. Posterior correlation plots for clade Ib model parameters.** (A) North and South Kivu; (B) Kamituga; (C) Other health zones of South Kivu; (D) Burundi. For each location,1,000 samples were drawn from the model-averaged posterior distribution. Panels in the upper triangular areas are 2-D histograms and those in the lower triangular areas are scatter plots. The notations $p$ and $q$ represent the proportion of high-activity individuals among males and females, respectively, and the brackets indicate age bins. The mean neighbour degrees $w_F$ and $w_M$ are normalised by the dominant eigenvalue of the community contact matrix $\lambda_C$.

##### ***Sensitivity analysis 1: synthetic contact matrix***

We used synthetic contact matrices for DRC and Burundi in place of the empirical contact matrices from Zimbabwe (Figure S6). The Watanabe-Akaike weights strongly preferred the contact matrix at home, effectively excluding the all-setting contact matrix as an alternative. The estimated fraction of the effective reproduction number attributable to sexual contact was 48% (95% CrI: 44–52%), 66% (61–71%), 22% (15–28%) and 21% (16–26%) for the Kivus, Kamituga, other health zones and Burundi, respectively. Higher estimates than those from empirical contact matrices may be because the model underestimated cases aged 20–29 than observed in the endemic provinces in 2024 (Figure S1F).

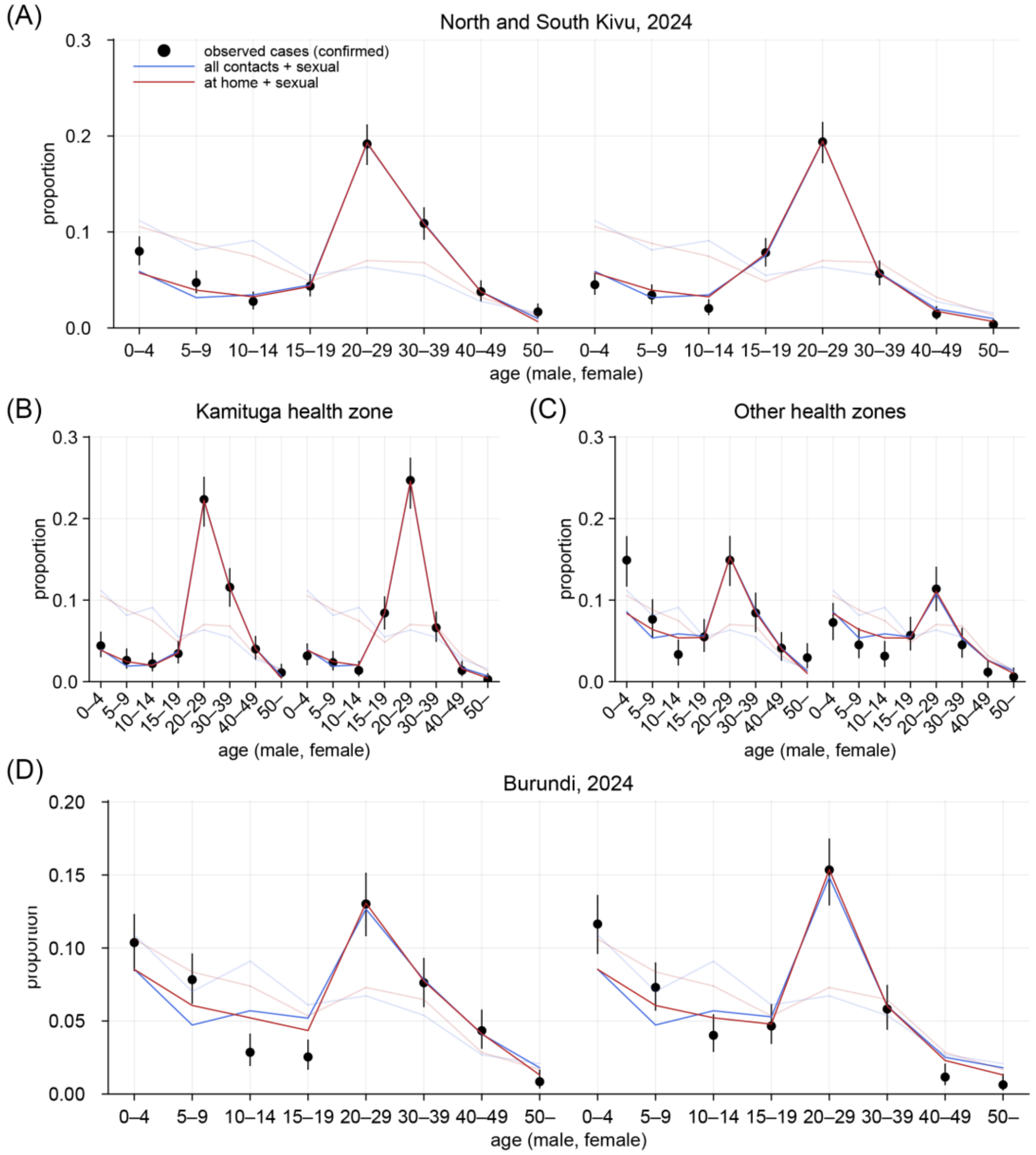


**Figure S6. Modelled age distributions of mpox clade I cases using synthetic matrices.** (A-D) Clade Ib cases from multiple settings in DRC and in Burundi, January–August 2024. Those aged 45 and older were assumed to be fully vaccinated in DRC and 70% vaccinated in Burundi. Half-transparent lines represent the model fit without assuming transmission over sexual contact.

##### *Sensitivity analysis 2: model assumptions*

We introduced a number of assumptions into our models, including but not limited to those on sexual contact patterns. As a sensitivity analysis, we re-estimated the proportion of $R_{\mathrm{eff}}$ attributable to sexual transmission, our main outcome of interest, in several alternative scenarios. Overall, the results suggested that our model was highly robust to these assumptions (Figure S7).

*(i) Proportion of high-activity individuals*

In the main analysis, we assumed that high-activity males account for 10% of those of sexually-active age and that high-activity females account for 1% of total female population. Instead of these baselines, we assumed 2-fold (for males) or 5-fold (for females) higher and lower proportions. Note that these changes scale $P$ or $Q$ in Equation 3 while do not affect $S_X$ or $\Sigma_X$ as the changes cancel out as in Equation 6.

*(ii) Age assortativity*

We assumed sexual contacts among high-activity groups are randomly assigned regardless of age (proportionate mixing). However, a previous study from South Africa[10] reported some age assortativity for casual sexual partners (Gupta's Q[11] of 0.357 among female and 0.393 among male participants). We inflated the diagonal elements of matrices $S_X$ or $\Sigma_X$ such that their Gupta's Q match the reported values:

$$s_{ij}^g = (1-g)s_{ij} + \delta_{ij} g \sum_k s_{kj}, \qquad (S1)$$

where $s_{ij}$ is the $(i,j)$-th entry of $S_X$ or $\Sigma_X$ (note that the direction of transmission is from $j$ to $i$), $g$ is the Gupta's Q and $\delta_{ij}$ is the Kronecker's delta. We can confirm that Gupta's Q of the diagonal-inflated matrix, defined as $\frac{1}{d-1}\sum_i \left(\frac{s_{ij}^g}{\sum_k s_{ki}^g} - \frac{\sum_j s_{ij}^g}{\sum_{ij} s_{ij}^g}\right)$ (where $d$ is the matrix dimension)[12], indeed becomes $g$ by noting that Q of the original proportionate mixing matrices is 0.

*(iii) Potential role of locality of lesion in transmission risks*

A recent study reported that adult cases (aged 15 or older) infected with clade Ib more frequently presented anogenital lesions than were typically observed among child cases or those infected with clade Ia[13], suggesting a possible association between the exposure route and lesion distribution. If such an association exists, transmission chains may favour a consistent contact route, i.e. transmission switching the contact routes (sexual contact transmission from community-acquired cases, or vice versa) may be less likely. To account for this hypothesis, in this scenario we reduced the risk of such "crossover" transmissions switching the contact routes by 10-fold. This is done by scaling down blocks corresponding to crossover transmission in the 4-by-4 block matrix in Equation 3:

$$\begin{bmatrix} O & O & S_{MF} & \phi\Sigma_{MF}Q \\ \phi C_{MM} & C_{MM} & \phi C_{MF} & C_{MF} \\ S_{FM} & \phi\Sigma_{FM}P & O & O \\ \phi C_{FM} & C_{FM} & \phi C_{FF} & C_{FF} \end{bmatrix}, \qquad (S2)$$

where $\varphi$ is a scaling factor of 0.1.

*(iv) Location-specific vaccine coverage*

Due to the limited geographically-disaggregated smallpox vaccine coverage data in DRC and Burundi, in the main analysis we assumed that the population-level protection conferred by smallpox vaccines across modelled locations in 2024 is uniform and identical to the estimate from clade Ia-endemic provinces. To address a possible regional variation in vaccination coverage, as well as potentially different vaccine effectiveness against clade Ib vs clade Ia, we allowed the effective vaccine coverage parameter $\varepsilon$ to vary across clade Ib-circulating locations. This yielded model averaging estimates for the relative susceptibility among smallpox-immunised cohorts of 0.33 (95% CrI: 0.22–0.47) for the Kivus, 0.35 (0.19–0.61) for Kamituga, 0.38 (0.23–0.58) for other health zones of South Kivu and 0.16 (0.09–0.25) for Burundi. These estimates overlap with the baseline estimate of 0.26 (0.17–0.38) and do not affect the estimated relative contribution of sexual contacts (Figure S7). Note that these individual estimates should be viewed with caution due to potential overfitting and lack of model validation (as opposed to our main model for clade Ia).

*(v) Alternative model weights*

The Watanabe-Akaike weights used for the model averaging in the main analysis was produced by combining only the WAIC values for the endemic provinces (both historical and recent outbreak datasets) and the Kivus, disregarding the WAIC values for the subprovincial datasets (i.e. Kamituiga and other health zones of South Kivu) or Burundi. Here we compared the model-averaged relative sexual contribution for two alternative weights. As the first set of alternative weights (weights 1), we replaced the WAIC for the Kivus with a mixture of the WAIC values of the Kivus, Kamituga and other health zones of South Kivu. Since the majority of the Kivus data consisted of cases from South Kivu, simply aggregating these three WAIC values would result in double counting; we thus downscaled the contribution of each WAIC by half. As the second set of alternative weights (weights 2), we further incorporated Burundi's WAIC into weights 1. The weight for a given model $M$ is then described as:

$$\begin{aligned}
&\omega_0(M) \propto \exp\left[{W_M}^{\mathrm{en1}} + {W_M}^{\mathrm{en2}} + {W_M}^{\mathrm{Kivus}}\right] \\
&\omega_1(M) \propto \exp\left[{W_M}^{\mathrm{en1}} + {W_M}^{\mathrm{en2}} + \frac{1}{2}\left({W_M}^{\mathrm{Kivus}} + {W_M}^{\mathrm{Kamituga}} + {W_M}^{\mathrm{OtherHZ}}\right)\right] \qquad (S3) \\
&\omega_2(M) \propto \exp\left[{W_M}^{\mathrm{en1}} + {W_M}^{\mathrm{en2}} + \frac{1}{2}\left({W_M}^{\mathrm{Kivus}} + {W_M}^{\mathrm{Kamituga}} + {W_M}^{\mathrm{OtherHZ}}\right) + {W_M}^{\mathrm{Burundi}}\right],
\end{aligned}$$

where $\omega_0$, $\omega_1$ and $\omega_3$ correspond to the baseline weights, weights 1 and weights 2, respectively. $W_M$'s are (-1/2) times the WAIC values for model $M$, whose superscript represent: the historical clade Ia data (en1; Dataset 2 of Extended Table 1), the recent clade Ia data (en2; Dataset 3), the Kivus (Dataset 4), Kamituga (Dataset 5), other health zones of South Kivu (Dataset 6) and Burundi (Dataset 7).

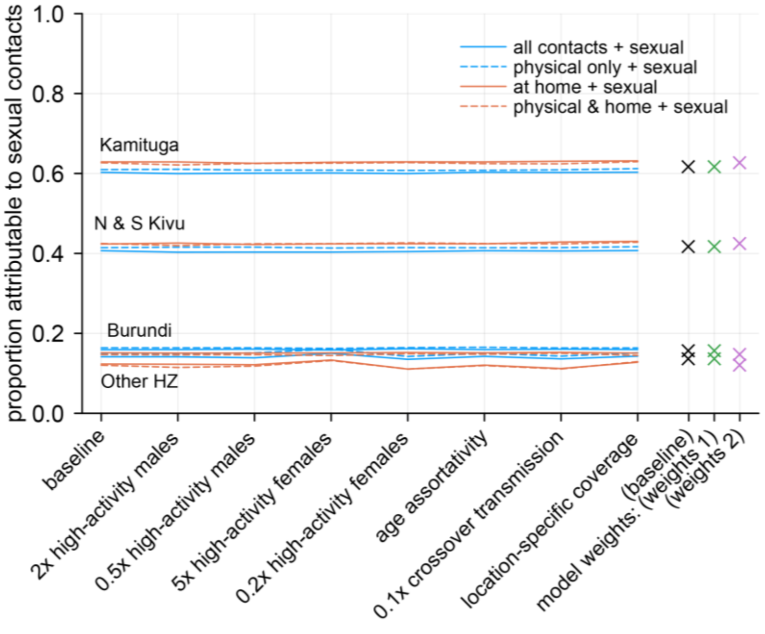


**Figure S7. Estimated relative contributions of sexual contacts to the overall transmission for sensitivity analysis scenarios.** Lines represent the median estimates of the proportion of $R_{\mathrm{eff}}$ attributable to sexual contacts in models using different contact matrix type. Cross marks represent the model-averaged estimates using Watanabe-Akaike weights based on different sets of data. The same set of colours and line types are used across four locations for simplicity—for the avoidance of confusion, the relative magnitudes between estimates are consistently Kamituga, North & South Kivu, Burundi and other health zones of South Kivu in descending order for any model or weights represented by the same colour / line type.

*(vi) Additional scenarios implicitly addressed: sexual transmission asymmetry and the ratio between mean degree and mean neighbour degree*

We describe two other scenarios of potential interest, which can be related to the scenarios already explored in Figure S7. First, we assumed that the transmission risk per contact does not vary by the directionality of transmission (male-to-female or female-to-male) in the main analysis, which may not always hold for general STIs[14–17]. If this assumption is violated, i.e. per-contact transmission risk from males to females is $\gamma$ times that from females to males, the contact block matrix becomes

$$\begin{bmatrix} O & O & S_{MF} & \Sigma_{MF}Q \\ C_{MM} & C_{MM} & C_{MF} & C_{MF} \\ \gamma S_{FM} & \gamma\Sigma_{FM}P & O & O \\ C_{FM} & C_{FM} & C_{FF} & C_{FF} \end{bmatrix}, \qquad (S4)$$

However, these changes could be absorbed by redefining $v_M$ and $w_M$ as $\gamma$ times each, except for the reciprocity requirement in Equation 7, which becomes

$$v_F \sum_a m_a q_a = \frac{v'_M}{\gamma} \sum_a n_a p_a, \qquad (S5)$$

where $v_M' = \gamma v_M$ is the redefined parameter, i.e. $\gamma$ times the mean sexual contact rate of males. This shows that the effect of this sexual transmission asymmetry is equivalent to scaling $p_a$ (proportion of high-activity males) $\gamma$ times, which we explored in the scenario (i).

Second, the ratio between $v_F$ and $w_F$ was specified at 2.17 using the reported number of paying and non-paying sexual partners among sex workers[18]. As a baseline assumption, we assumed that the numbers of paying and non-paying partners are uncorrelated. However, it is plausible that these numbers are correlated, leading to an increase in this ratio. In the most extreme scenario where the numbers are completely correlated (i.e. the covariance becomes the product of the standard deviations of the number of clients and the number of non-paying partners), this ratio becomes 3.16. Using this ratio to specify $v_F$ and then $v_M$ through reciprocity leads to a 1.5-fold reduction in both of these parameters. This change is equivalent to using 1.5-fold lower proportions of high-activity groups ($p_a$ and $q_a$), a smaller deviation from the baseline than explored in the scenario (i).

***Speed of convergence to eigenvector***

We assessed the validity of our assumption that the observed distributions of cases are representative of the dominant eigenvector of the next generation matrix. First, we used the age distribution of cases with zoonotic exposure in the 2011–2015 Tshuapa dataset (i.e. Dataset 1 in Extended Data Table 1) as the initial distribution (generation 0). We then iteratively left-multiplied this distribution with the next generation matrix based on each type of contact matrix to generate the distribution of subsequent human-to-human transmission generations. The results showed that the convergence is rapid enough for all human-exposed cases (generation 1 onward) to follow the dominant eigenvector (Figures S8A). Similarly, for our model of a mixed mode of community and sexual contact transmission, we simulated the age-sex distribution of cases by generations starting from high-activity initial cases (all in the age group 20–29 and evenly distributed between males and females). The age-sex distribution exhibited a rapid convergence (Figures S8B).

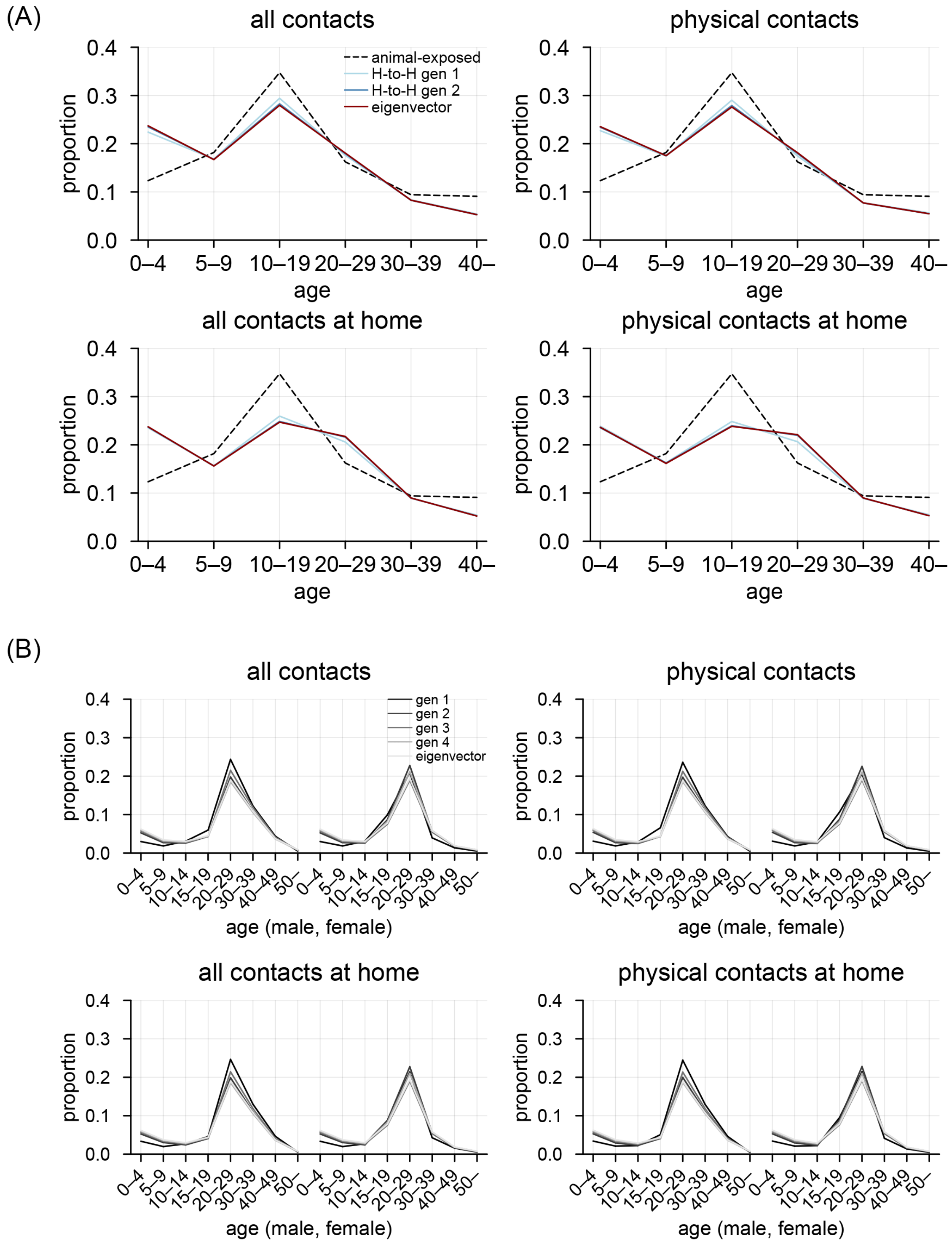


**Figure S8. Convergence of case distribution to the dominant eigenvector of the next generation matrix.** (A) Simulated age distributions of cases across generations based on different contact matrices with community contact transmission only. Initial cases were assumed to follow the reported age distribution of cases with zoonotic exposure in Tshuapa, 2011–2015. (B) Simulated age and sex distributions of cases across generations based on different contact matrices with both community and sexual contact transmission. Initial cases were assumed to be all 20–29 and evenly distributed between males and females.

***Outbreak potential of clade I among MSM in a population with immunity from the previous clade IIb global outbreak***

In our main analysis, we only focused on the heterosexual contact network and did not consider transmission over the men who have sex with men (MSM) sexual contact network. As MSM have been the most affected group in the clade IIb global outbreak since 2022, assessment of clade Ib's transmission potential in this group, particularly in the presence of immunity, is of public health interest.

To assess the outbreak potential of clade I in a population with infection-derived and/or vaccine-derived immunity from the previous clade IIb outbreak and vaccination, we used a deterministic transmission model accounting for highly heterogeneous distribution of sexual partners among MSM developed in Murayama et al.[19], which we updated using a 4-week partnership data (as opposed to the 1-year partner data used in the original study) from the British National Surveys of Sexual Attitudes and Lifestyles[20,21]. We assumed that an MSM population that previously experienced a clade IIb epidemic had reached the natural final epidemic size and established long-term herd immunity. The initial effective reproduction number of a new clade in such a population with assumed completely cross-protective immunity from the clade IIb epidemic is then given as:

$$R_{\mathrm{eff}}^{\mathrm{new}} = \frac{\beta_{\mathrm{new}} \int_1^{\infty} x(x-1) S_{\mathrm{final}}(x) dx}{\langle x \rangle}, \qquad (S6)$$

where $\beta_{\mathrm{new}}$ is the secondary attack risk (SAR) per sexual partnership for the new clade, $\langle x \rangle$ is the mean degree (the average number of sexual partners) and $S_{\mathrm{final}}$ is the number of susceptible individuals with $x$ partners at the end of the previous epidemic divided by the total population size[19]. Note that the bracket is the expectation operator over the population. For a clade I major outbreak to occur, the initial effective reproduction number needs to meet the condition of $R_{\mathrm{eff}}^{\mathrm{new}} \geq 1$, which gives us the threshold for $\beta_{\mathrm{new}}$ and thus the basic reproduction number ($R_0^{\mathrm{new}}$) required for the epidemic takeoff. In the presence of population-level vaccine protection, the required $R_0$ would be $1/(1-\nu)$-fold higher, where $\nu$ is the relative risk of infection among the vaccinated population. We assumed vaccines were randomly allocated and with an effectiveness of 86%[22]. For simplicity, we neglected the impact of vaccination on the real-time dynamics of the previous clade IIb outbreak was minimal, as suggested by previous studies[23,24]. We defined the effective susceptible proportion as the ratio between the initial effective reproduction number of a new clade (Equation 10) and its basic reproduction number in a fully-susceptible population:

$$S_{\mathrm{eff}} = \frac{R_{\mathrm{eff}}^{\mathrm{new}}}{R_0^{\mathrm{new}}} = \frac{\int_1^{\infty} x(x-1) S_{\mathrm{final}}(x) dx}{\langle x(x-1) \rangle}. \qquad (S7)$$

Note that $R_0^{\mathrm{new}} = \beta_{\mathrm{new}} \langle x(x-1) \rangle / \langle x \rangle$.

By simulating the model, we showed the effective susceptible proportion and $R_0$ required for clade I's epidemic takeoff as a function of $R_0$ of clade II in the previous outbreak (Figure S9). $R_0$ for clade IIb has been estimated mostly around 2 to 3[25,26,24]. Within this range, the effective susceptible proportion is 10%-30%, requiring an $R_0^{\mathrm{new}}$ of 3 to 8 to cause a major outbreak even without any vaccination. If 50% of the population is vaccinated, the effective susceptible proportion drops to about 5%-15%, requiring an $R_0^{\mathrm{new}}$ of at least 6.

These results suggest that the risk of another major mpox outbreak within MSM populations may be low in countries affected by the 2022 outbreak if existing immunity confers sufficient cross-protection. However, they should be interpreted with caution due to the simplistic nature of the model and uncertainties associated with e.g. sexual behaviour data, impact of behavioural changes and potential waning of immunity.

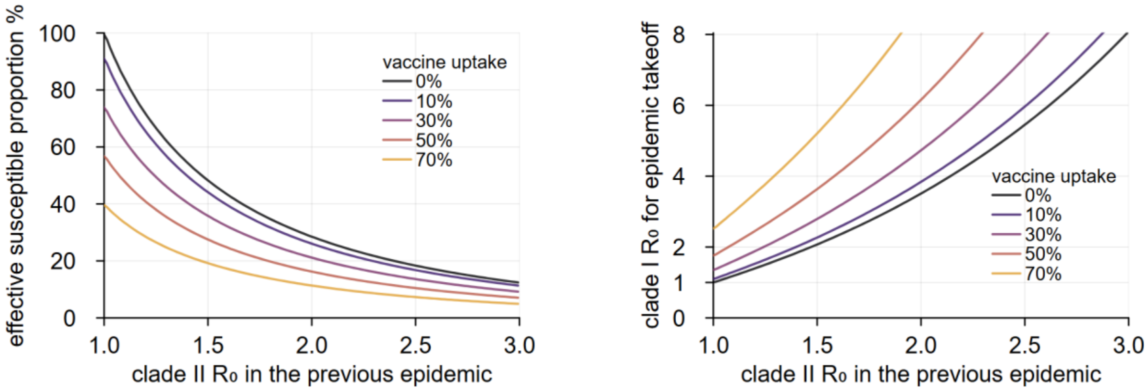


**Figure S9. Outbreak potential of clade I within MSM community with herd immunity established in the clade IIb global outbreak.** The relationships between the basic reproduction number for clade IIb and (A) effective susceptible proportion represented as $R_{\mathrm{eff}}^{\mathrm{new}}/R_0^{\mathrm{new}}$ and (B) the clade I's basic reproduction number required to initiate a major outbreak with different vaccination uptake.

##### *Proof of concept using an individual-level network model*

In our next generation matrix model, we represented the heterogeneous sexual partnership network by two classes: high- and low-sexual activity populations. Despite this simplification of highly-heterogeneous sexual behaviours (typically characterised by heavy-tailed distributions) in mpox and other sexually-transmitted infections[27–29], its impact on the conclusions is likely minimal as our modelling scope was the initial exponential growth of mpox. Here we provide a proof of concept using a simple individual-level network model. To focus on modelling of individual-level heterogeneity in sexual partnerships, age-dependent mixing or differential susceptibility was not considered in this analysis for simplicity.

As in the main analysis, we considered networks where links represent community or sexual contacts. Here we consider networks consisting of 5,000 male nodes and 5,000 female nodes, of which 500 (10%) and 100 (2%) are assigned to high-activity groups, respectively, roughly in line with the proportions of high-activity individuals among the sexually-active age groups (aged 15–49) in the main analysis. We constructed two versions of networks assuming different numbers of high-activity sexual connections (networks 1 and 2; see Table S2). For each version, we first drew community contact links between these nodes by the Erdos-Renyi algorithms[30] with a mean degree of 7.5. We then drew bipartite sexual contact links (i.e. exclusively between opposite sexes) to form a mixture of scale-free and Erdos-Renyi networks in the following steps. Scale-free networks with a heavy-tail (power-law) degree distribution were assumed to represent sexual connections within high-activity groups (high-activity sexual network); Erdos-Renyi networks are less heterogeneous and are assumed to connect through all individuals regardless of sexual activity (low-activity sexual network).

1. For the high-activity sexual network, first generate two scale-free networks of an exponent parameter of 3 with 500 nodes each (i.e. for males and for females) using Cho et al.'s method[31]. The number of links in each network was set such that the mean degree becomes as specified in Table S2 (i.e. 1,000 links in network 1 and 500 links in network 2).

2. Randomly pair up a link from the male scale-free network and another link from the female scale-free network, thereby generating pairs of a male-to-male link and a female-to-female link. For each of the pairs (involving two male and two female nodes), dissolve the two existing same-sex links and instead (randomly) draw two new opposite-sex links such that each node has one opposite-sex neighbour. This creates a bipartile network of 1,000 nodes (500 males and 500 females).
3. Reduce the number of female nodes to 100 by merging every five female nodes into one. Each group of five are chosen in the order of node indices $i$'s in Cho et al.'s method (i.e. those with similar degrees). Parallel links are simplified if any. The resulting bipartile network has 500 male and 100 female nodes, which we use as the high-activity sexual network.
4. For the low-activity sexual network, first generate two Erdos-Renyi networks with 5,000 nodes each and a mean degree of 0.5. Transform these two networks into one bipartile network of 10,000 nodes in the same way as the step 2.
5. Merge the high-activity and low-activity sexual networks by associating the first 500 male and 100 female nodes in the low-activity sexual network with the high-activity male and female nodes; i.e. the 500 male and 100 female high-activity nodes inherit sexual links from both high-activity (scale-free) and low-activity (Erdos-Renyi) networks, whereas the remaining low-activity nodes only have Erdos-Renyi sexual links.

To capture the stochastic fluctuations, we repeated the above network construction process to obtain 100 realisations each for networks 1 and 2. Figures S10A and S10B illustrate the network structures, where the number of nodes and links are scaled down by 10 folds for visualisation.

For complex networks, the dominant eigenvalue of the adjacency matrix of a network is known to define the basic reproduction number $R_0$[32]. The relative contribution of sexual contacts to $R_0$ for networks 1 and 2, measured as the relative reduction in the dominant eigenvalue of adjacency matrix when all sexual contact links are removed, generally aligned with the range of our estimates for clade Ib in the main analysis (Table S2). Using these networks, we tested our next generation matrix approach with two sexual activity classes by comparing the dominant eigenvalues between the adjacency matrix and the next generation matrix. The next generation matrix assuming no age-dependent mixing or differential susceptibility can be obtained by adapting Equation 3 where the block matrices accounting for age groups are replaced with scalars:

$$\begin{bmatrix} O & O & s_F & \sigma_F q \\ c & c & c & c \\ s_M & \sigma_M p & O & O \\ c & c & c & c \end{bmatrix}, \qquad (S8)$$

where $s_X$ is the mean neighbour degrees (i.e. degree-weighted means of degrees) and $\sigma_X$ is the mean degrees for sexual contact links among high-activity group of sex $X$. The entries of the 2nd and 4th rows, $c$, are half the mean degree for the entire network combining both community and sexual contact links; this is to be consistent with our use of contact matrices divided in half in the main analysis, where the reported contacts included sexual contacts by definition. The proportions of high-activity males ($p$) and females ($q$) were respectively set at 0.1 and 0.02, as were assumed in the network construction.

Comparison of the dominant eigenvalues of the adjacency and next generation matrices for each realisation of networks suggests that the approximation by the next generation matrix with two classes has a reasonable performance (Figure S10C).

**Table S2. Characteristics of proof-of-concept networks.**

| | | Network 1 | Network 2 |
|---|---|---|---|
| Mean degree | Community contacts | 7.5 | 7.5 |
| | Sexual contacts (high-activity group) | Male: 4.5<br>Female: 20.5 | Male: 2.5<br>Female: 10.5 |
| | Sexual contacts (low-activity group) | 0.5 | 0.5 |
| Dominant eigenvalue (median [95% range]) | | 15.6 [15.1–16.1] | 10.8 [10.4–11.2] |
| % $R_0$ attributable to sexual contacts (median [95% range]) | | 42% [40–43%] | 15% [13–19%] |

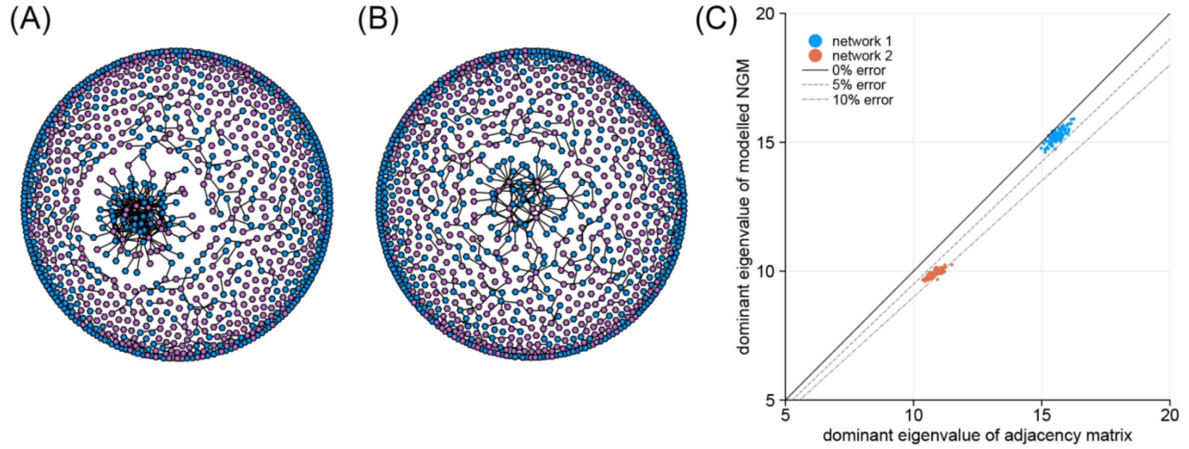


**Figure S10. Next generation matrix approximation for proof-of-concept network models.** Illustration of sexual contact networks for (A) network 1 with a higher connectivity within the high-activity sexual network and (B) network l with a lower connectivity within the high-activity sexual network. Blue and purple nodes represent male and female, respectively. The networks are generated using the same algorithm as used for the analysis but with 10 times fewer nodes and links for visualisation. Community contact links are omitted from the visualisation. (C) Comparison between the dominant eigenvalue of the adjacency matrix of the proof-of-concept networks and the dominant eigenvalue of the modelled next generation matrix. Each dot represents one of the 100 generated networks.

***Proof of concept with degree assortativity***

The previous section suggested that our simple next generation matrix approach can approximate the transmission dynamics in a complex network consisting of sexual and community contacts whose sexual contact element is a configuration network assuming no degree assortativity. This assumption of no degree assortativity allowed us to specify $s_X$ as the mean neighbour degree in the proof-of-concept model. While this may no longer be appropriate in the presence of degree assortativity, our next generation matrix approach could still be applicable with alternative values for $s_X$. We constructed an alternative next generation matrix by substituting $s_F$ and $s_M$ in Equation S8 with $\lambda_F$ and $\lambda_M$, respectively, by which we denote the weighted mean degrees among female and male nodes using the eigenvector centrality[33,34] as weights. In a network with little or no degree assortativity, $s_X$ and $\lambda_X$ are known to be similar[34]. The alternative next generation matrix therefore provides equivalent (or slightly improved) performance in capturing the dominant eigenvalue of the adjacency matrix for the original networks with no sexual contact degree assortativity (Figure S11A). We then applied these two next generation matrices to networks with increased sexual contact degree assortativity: we followed the same network generation process described in the previous section, except that in step 2 where two new opposite-sex links are drawn, we linked them in a degree-assortative manner (i.e. paired the higher-degree male with the higher-degree female, and the lower-degree male with the lower-degree female) rather than randomly. This led to the resulting networks to have a (directed) degree assortativity[35] of about 0.15 (95 percentile: 0.1–0.2). The performance of the original next generation matrix deteriorated for these assortative networks (as $s_X$ could no longer approximate $\lambda_X$), whereas that of the alternative matrix remained robust (Figure S11B). These results suggest that our modelling approach might still be useful in the presence of degree assortativity, although with a caveat of simplifying assumptions including disregarded age structures in the proof-of-concept model. That is, even if degree assortativity exists in the proof-of-concept network, the next generation matrix in Equation S8 can still be adapted to capture it by only replacing $s_X$ (mean neighbour degrees) with $\lambda_X$. In the main analysis, $w_X$ (scaling for $S_X$, which is analogous to $s_X$) were estimated as free parameters, which suggests that our model had a similar flexibility to capture the dynamics even in the presence of certain degree assortativity.

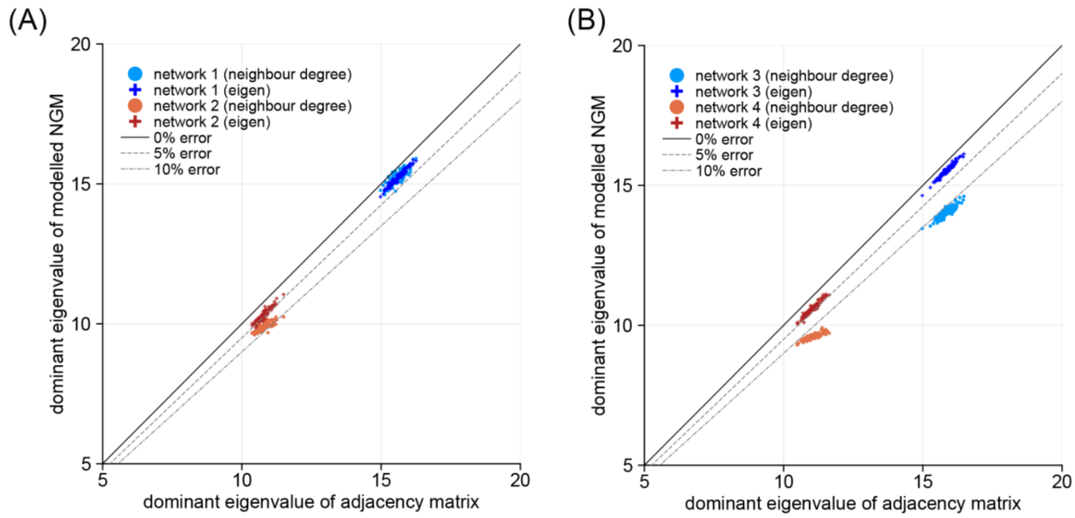


**Figure S11. Next generation matrix approximation for proof-of-concept network models with and without degree assortativity.** Comparison between the dominant eigenvalue of the adjacency matrix of the proof-of-concept networks and the dominant eigenvalue of the modelled next generation matrix: (A) original networks with no degree assortativity and (B) networks constructed to have a degree assortativity. Circles represent the next generation matrix using the mean neighbour degree as were shown in Figure S10C and crosses represent the alternative next generation matrix using the eigenvector centrality-weighted mean degree. Each dot represents one of the 100 generated networks.

#### Comparison of characteristics between DRC, Burundi and Zimbabwe

To construct community contact matrices in the main analysis, we used empirical contact data from Zimbabwe as a proxy for DRC and Burundi. As an assessment of this assumption of similarity in contact behaviour between these countries, we compared their country characteristics selected in Prem et al.[36] as potentially relevant to contact patterns using principal component analysis (PCA)[37]. A total of 75 country characteristics variables (e.g. age/sex distributions, population in urban and rural settings, gross domestic product per capita, internet use, tuberculosis incidence) from 115 countries for which those variables were available were included in the analysis[38–43].

The first four principal components (PCs) explained 72.3% of the total data variance. PC1 generally consisted of variables related to population age structures, longevity/fertility and infrastructure. PC2 was associated with working age population in general workplaces and schools. PC3 consisted of variables related to working age populations at schools, government expenditure on education, unemployment rates, population density and HIV prevalence. PC4 was characterised by adult population structures, net migration rate, agricultural land use and urban-rural population ratios.

The PCA results suggested that countries mapped over those PCs clustered within geographical regions (Figure S12). Within Africa, DRC, Burundi and Zimbabwe are particularly in close proximity, suggesting that they overall share similar characteristics.

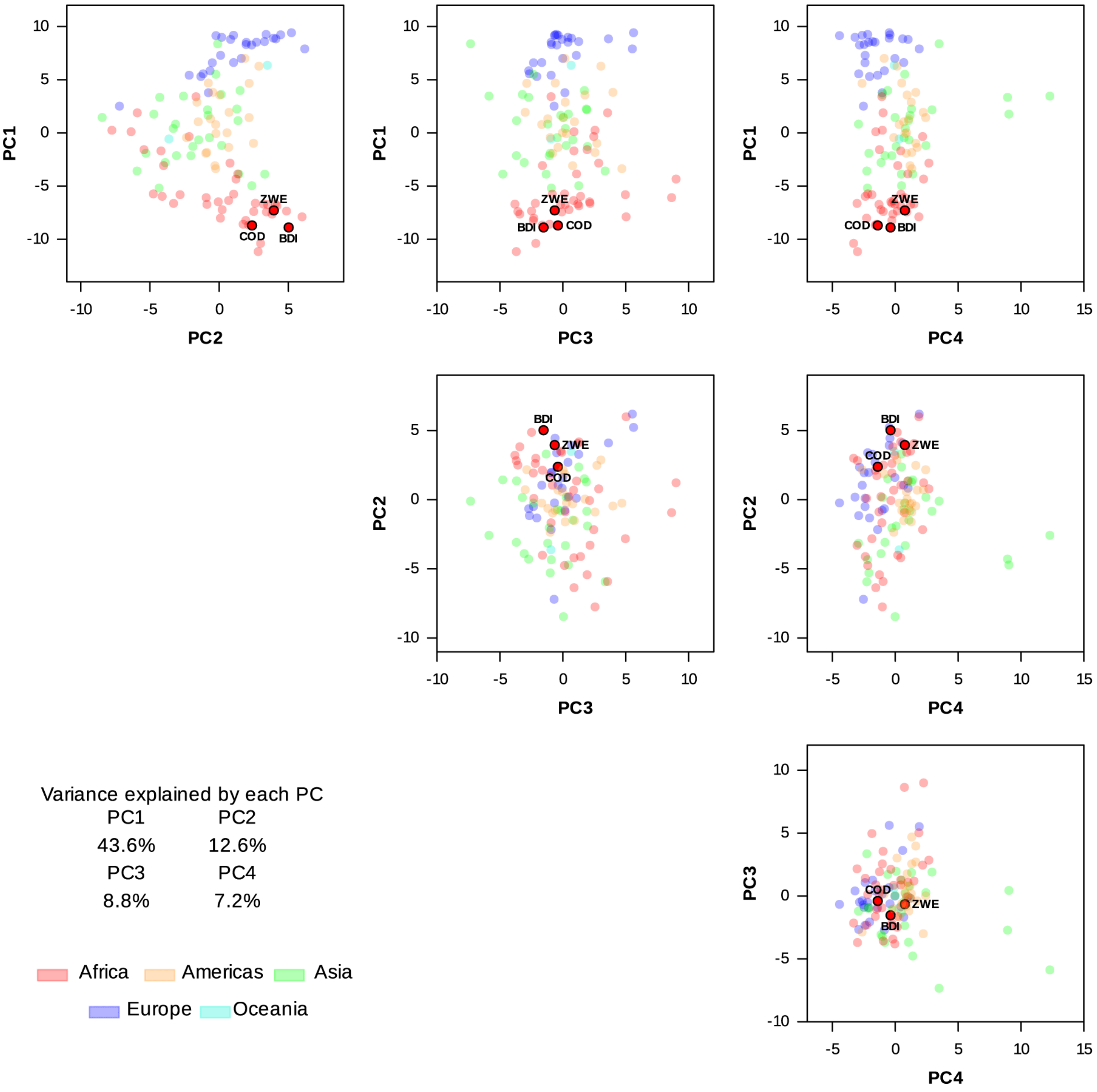


**Figure S12. Principal component plots comparing country-specific characteristics for DRC, Burundi, Zimbabwe and 112 other countries.** Each dot corresponds to one of the included countries, of which DRC ("COD"), Burundi ("BDI") and Zimbabwe ("ZWE") are marked with black borders.

38. World Population Prospects. https://population.un.org/wpp/.

39. UNdata | record view | Population by age, sex and urban/rural residence. https://data.un.org/Data.aspx?d=POP&f=tableCode%3A22.

40. ILO Modelled Estimates (ILOEST database). *ILOSTAT* https://ilostat.ilo.org/methods/concepts-and-definitions/ilo-modelled-estimates/.

41. Home - UIS Data Browser. https://uis-data-browser-frontend-enmhah4ay-ixt1.vercel.app/index.

42. Teachers by age. *OECD* https://www.oecd.org/en/data/indicators/teachers-by-age.html.

43. Class size and student-teacher ratios. *OECD* https://www.oecd.org/en/topics/class-size-and-student-teacher-ratios.html.
